# Understanding the robustness of vision-language models to medical image artefacts

**DOI:** 10.1101/2025.05.13.25327495

**Authors:** Zijie Cheng, Ariel Yuhan Ong, Siegfried K. Wagner, David A. Merle, Lie Ju, Boxuan Li, Tiantian He, An Ran Ran, Hongyang Jiang, Dawei Yang, Ke Zou, Jocelyn Hui Lin Goh, Sahana Srinivasan, Andre Altmann, Daniel C. Alexander, Carol Y. Cheung, Yih Chung Tham, Pearse A. Keane, Yukun Zhou

**Affiliations:** Department of Medical Physics & Biomedical Engineering, University College London, London, UK; Institute of Ophthalmology, University College London, London, UK; NIHR Biomedical Research Centre, Moorfields Eye Hospital NHS Foundation Trust, London, UK; Department of Ophthalmology and Visual Sciences, The Chinese University of Hong Kong, Hong Kong Special Administrative Region, China; Centre for Innovation and Precision Eye Health; and Department of Ophthalmology, Yong Loo Lin School of Medicine, National University of Singapore, Singapore; Ophthalmology and Visual Science Academic Clinical Program, Duke-NUS Medical School, Singapore, Singapore; Singapore Eye Research Institute, Singapore National Eye Centre, Singapore; The UCL Hawkes Institute, University College London, London, UK; Department of Computer Science, University College London, London, UK

**Keywords:** Vision-language models, image artefacts, model robustness, disease detection

## Abstract

Vision-language models (VLMs) can answer clinically relevant questions with their reasoning capabilities and user-friendly interfaces. However, their robustness to commonly existing medical image artefacts has not been explored, leaving major concerns in trustworthy clinical decision-making. In this study, we assessed the robustness of recent VLMs to medical image artefacts in disease detection across three different medical fields. Specifically, we included five categories of image artefacts, and evaluated the VLMs’ performance on images with and without artefacts. We build evaluation benchmarks in brain MRI, Chest X-ray, and retinal images, involving four real-world medical datasets. Our results demonstrate that VLMs showed poor performance on original unaltered images and performed even worse when weak artefacts were introduced. The strong artefacts were barely detected by those VLMs. Our findings indicate that VLMs are not yet capable of performing medical tasks with image artefacts, underscoring the critical need to explicitly incorporate artefact-aware method design and robustness tests into VLM development.

## Introduction

Vision-language models (VLMs) are trained using vast and diverse images paired with textual descriptions (prompts), allowing them to deeply understand visual content and generate accurate responses^1^. Powerful examples, such as GPT-4o^2^ and Claude 3.5 Sonnet^3^, enable diverse applications such as image captioning^4^. The advances have been extended to the medical field, leading to specialized medical VLMs such as MedDR^5^, LLaVAMed^6^, and BiomedCLIP^7^, which contribute to disease detection in multiple medical fields. However, in real-world medical scenarios, medical image artefacts are common and potentially bias the performance of VLMs, and this has not been systematically studied.

Image artefacts in the medical domain are distortions caused by patient movement, equipment limitations, technicians’ skills and environmental factors^8–12^. They are commonly seen in many medical imaging modalities such as magnetic resonance imaging (MRI)^13^, optical coherence tomography (OCT)^14^, and X-rays^15^. For instance, random motion in OCT imaging refers to movements of the subject, eye drift or microsaccades during scan acquisition^14,16,17^. These image artefacts can lead to errors in disease detection, including false positives, where normal cases are incorrectly identified as diseased, and false negatives, where diseased cases are mistakenly classified as normal^18–21^. These errors occur when image artefacts mimic pathological features or obscure actual abnormalities, respectively. Previous literatures have studied the impact of medical image artefacts on traditional AI models and have observed remarkable performance drops^22–24^.

Recent studies have explored the use of VLMs in medical visual question answering (VQA) to evaluate their capabilities in disease detection^25–29^. Their results show that some powerful VLMs (e.g., GPT-4o) could achieve performance comparable to junior doctors^30–34^ in certain tasks. However, few studies have explored the robustness and detection capabilities of VLMs when medical image artefacts are present in these tasks. Understanding this critical knowledge is essential for advancing trustworthy medical AI applications in clinical settings, especially as VLMs are increasingly seen as promising solutions due to their strong capabilities in reasoning and user interaction. Fig. 1 illustrates representative failure cases in disease detection by VLMs, attributable to the presence of medical image artefacts.

**Fig. 1:**
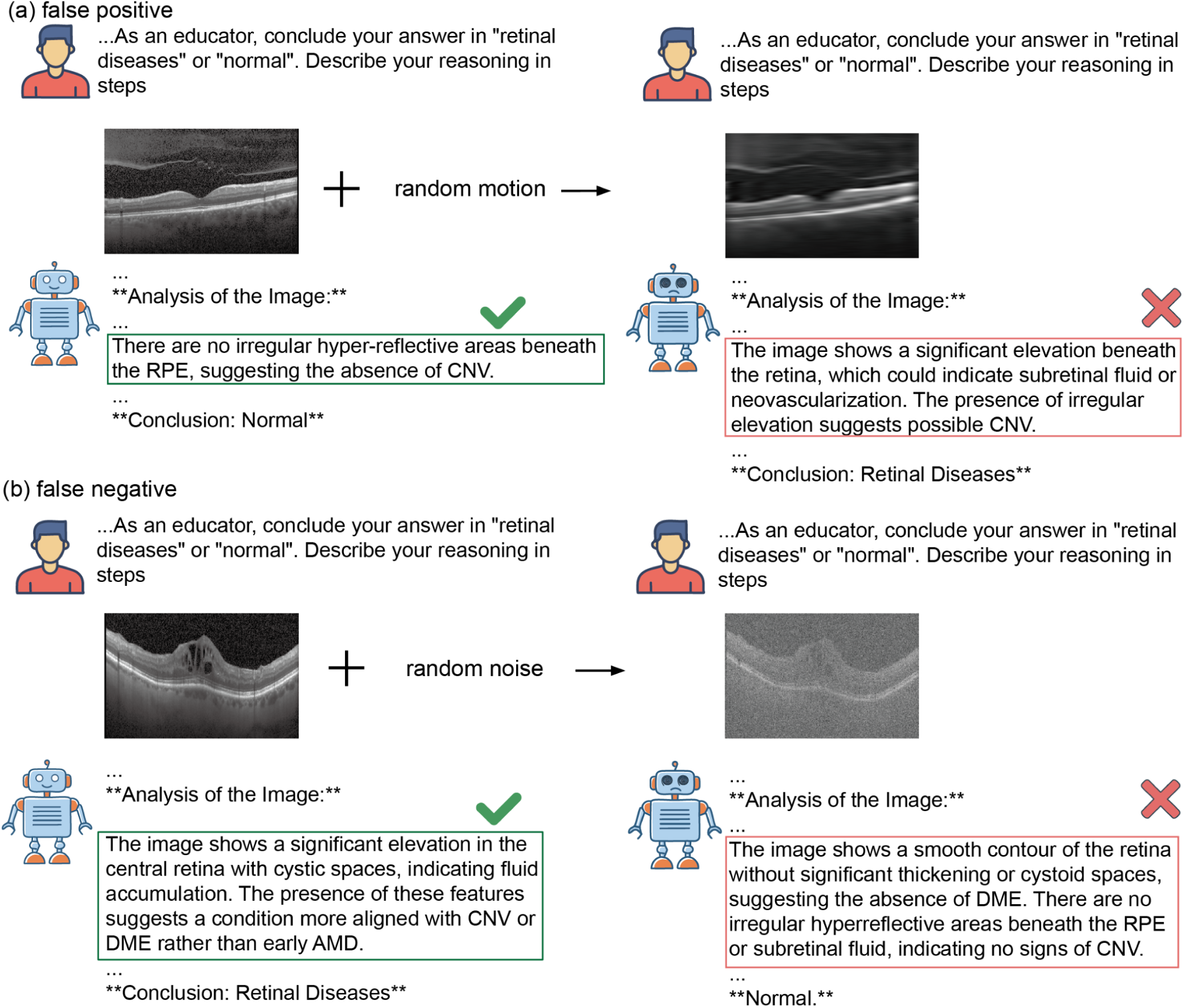
False positive and negative examples after adding artefacts to images. Figure (a), an example of a false positive case. Vision-Language models (VLMs) correctly classify normal cases when analysing original unaltered images, but misclassify the same cases as diseased when random motion is introduced. Figure (b), an example of a false negative case. VLMs correctly identify abnormal cases in original unaltered images, but misclassify them as normal after random noise is introduced. More misdetection examples with full responses from VLMs shown in Supplementary Table 1.

In this study, we summarise our contributions in three points: 1) We evaluated VLMs’ robustness in disease detection to medical image artefacts at different scales. To achieve that, 2) we constructed a benchmark spanning three medical fields by adding five different image artefacts and proposed three metrics to assess VLMs’ performance. The metrics include: model performance (i.e., accuracy and sensitivity), performance percentage change, which describe the proportional change in VLMs’ performance when weak artefacts are introduced to images, and the strong artefact detection rate, which measures the VLMs’ ability to identify images with strong artefacts. We also assessed their performance in detecting diabetic retinopathy from color fundus photographs with real-world artefacts to better demonstrate their robustness under clinical settings. 3) We revealed the insufficient robustness of current VLMs to medical image artefacts and underscore the urgent need to develop more robust medical VLMs for healthcare applications.

## Results

Figure 2 shows the overview of the project, including the construction of benchmarks and evaluation of VLMs’ robustness to medical image artefacts, spanning over three medical fields. For each medical field, we randomly selected 200 images, 100 for normal and 100 for disease cases. We built the benchmark by artificially introducing artefacts such as intensity artefacts (bias field, motion, and noise) and spatial artefacts (cropping and rotation) to the original unaltered images at increasing scales. More details can be found in the Benchmark subsection of the Method, Extended Figures 1∼3, and Supplementary Tables 2 and 3. This study included two proprietary models (GPT-4o and Claude 3.5 Sonnet) and two open-source models (Llama 3.2 and BiomedCLIP), with BiomedCLIP specifically designed for medical applications. We evaluated their performance on disease detection respectively with original unaltered images and images with weak artefacts. Additionally, we tested the VLMs’ capabilities in detecting the strong artefacts (further details are in the Metrics for model robustness and Models evaluated subsections of the Method section). We also assessed the impact of prompt engineering on VLMs’ performance from restricting reasoning to encourage step-by-step thinking, using structured, standard, and Chain of Thought prompts. P-values were calculated using a two-sided t-test to compare the original performance with the performance under weak artefacts for each task, to assess significance.

**Fig. 2:**
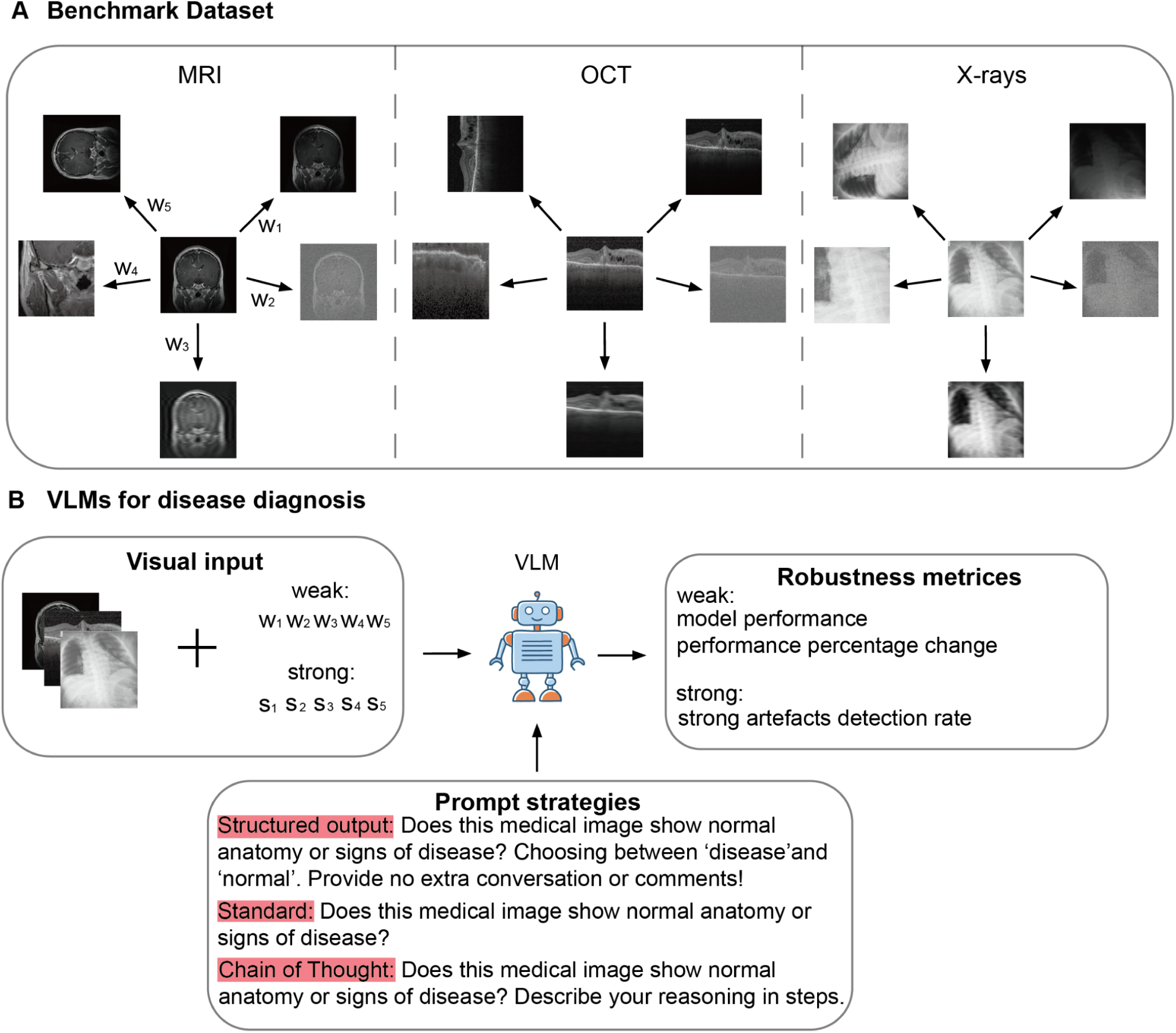
Overview of building benchmarks and the pipeline for evaluating robustness of Vision-Language models (VLMs) on disease detection tasks. Figure A, robustness evaluation benchmarks include three disease detection tasks corresponding to different image modalities. Three intensity artefacts (random bias field, noise, and motion) with weak (w₁∼w₃) and strong (s₁∼s₃) scales, and two spatial artefacts (random cropping and rotation) with weak (w₄∼w₅) and strong (s₄∼s₅) scales are introduced to original unaltered images. Figure B, project pipeline demonstrating how we evaluated the VLMs’ robustness. When adding weak artefacts, we evaluated model performance (e.g., accuracy) and the performance drop from its performance on the original unaltered images. When images were severely distorted by strong artefacts, we assessed their ability to detect poor image quality. All experiments were repeated through different prompt strategies from restricting reasoning through structured output to encouraging reasoning step by step through Chain of Thought.

### Model performance in original unaltered images

We assessed the performance of various VLMs on original unaltered images (Fig. 3). For brain tumour (MRI) and COVID-19/pneumonia detection (chest X-ray), BiomedCLIP achieved the highest accuracy, with scores of 0.770 (95% CI: 0.720 to 0.820) and 0.760 (95% CI: 0.710 to 0.805) respectively, using the standard prompt. This advantage also extended to sensitivity, particularly in MRI applications, where it reached the highest value of 0.950 (95% CI: 0.910 to 0.990) (Extended Data Fig. 4). In macular disease detection (OCT), GPT-4o achieved the highest accuracy of 0.778 (95% CI: 0.720 to 0.830) and sensitivity of 0.656 (95% CI: 0.560 to 0.740) using the standard prompt. In contrast, Llama 3.2 11B classified most cases as diseased, even normal ones, resulting in poor accuracy below 0.6, high sensitivity and low specificity across all tasks (Extended Data Figs. 4, 5).

**Fig. 3:**
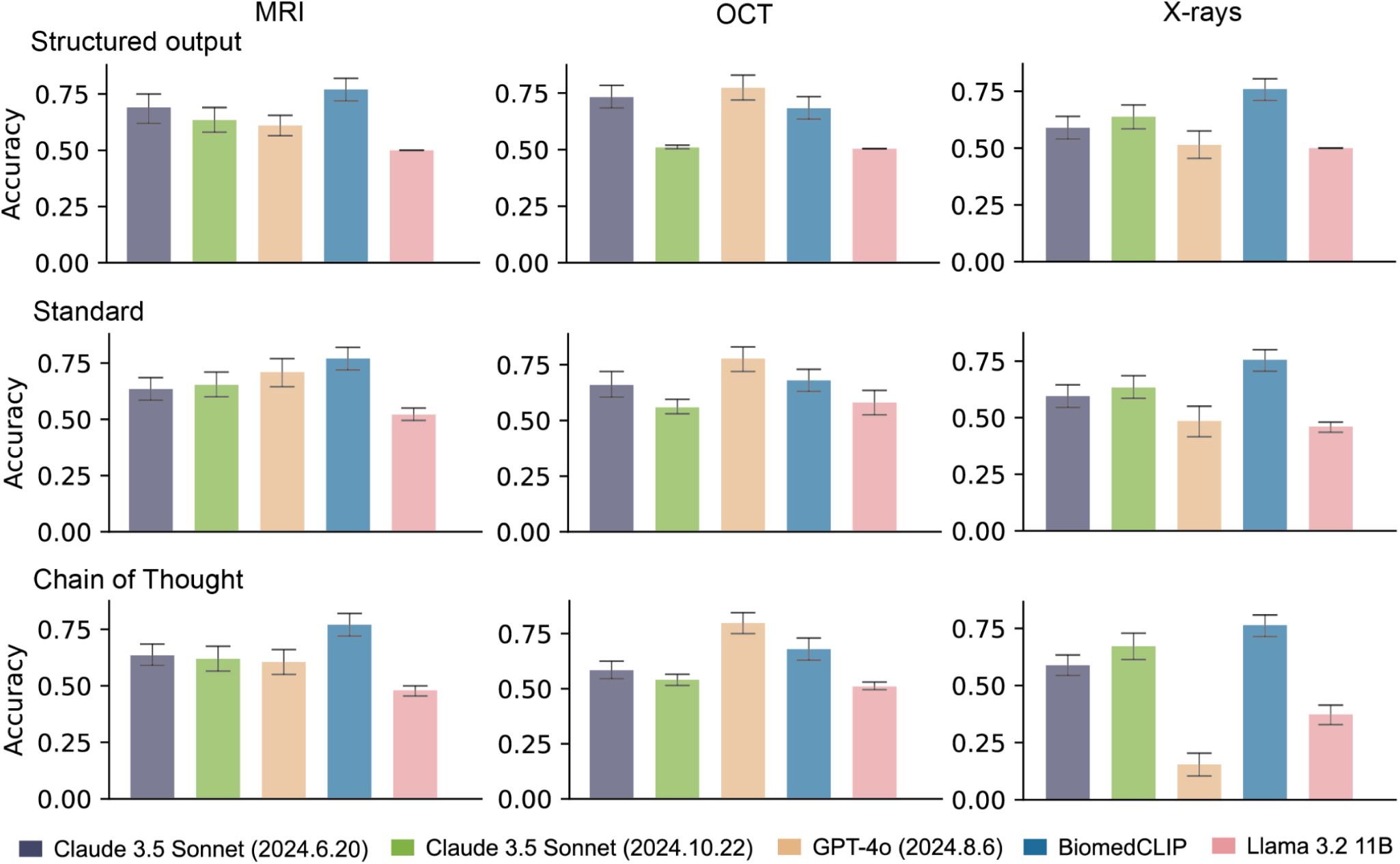
The accuracy of Vision-Language models (VLMs) in detecting disease on original unaltered images. BiomedCLIP requires only task categories and outputs each category probability, but we include it across all prompt strategies for comparison. Quantitative results are detailed in Supplementary Table 6. For each task, accuracy measurements derive from 1000 stratified bootstrap samples. Results show mean accuracy with 95% confidence intervals (error bars).

### Model performance on images with weak artefacts

We evaluated VLMs’ performance in disease detection using images with weak artefacts and compared it to original performance. We presented the lesion detection accuracy on images with weak artefacts for the top three models (Fig. 4a). These models were selected based on their performance on the original unaltered images using standard prompts (Fig. 3 and Extended Data Figs. 4 and 5). Among the models evaluated, BiomedCLIP demonstrated the highest accuracy and sensitivity in MRI applications. For instance, it achieved an accuracy of 0.801 (95% CI: 0.745 to 0.855) and sensitivity of 0.782 (95% CI: 0.700 to 0.860) when detecting images with weak random noise using the standard prompt (Extended Data Fig. 6). However, this advantage was not observed in other disease detection tasks. GPT-4o demonstrated the strongest performance in OCT applications. In particular, it outperformed BiomedCLIP in sensitivity, achieving 0.766 (95% CI: 0.705 to 0.825) for detecting macular diseases in images with weak random motion, compared to BiomedCLIP’s 0.120 (95% CI: 0.060 to 0.190) (Extended Data Fig. 7). In chest X-ray applications, no model showed clear superiority in both accuracy and sensitivity. For example, Claude 3.5 Sonnet (2024.10.22) achieved the highest sensitivity of 0.930 (95% CI: 0.870 to 0.980), but its accuracy was moderate at 0.505 (95% CI: 0.470 to 0.540) (Extended Data Fig. 8).

**Fig. 4:**
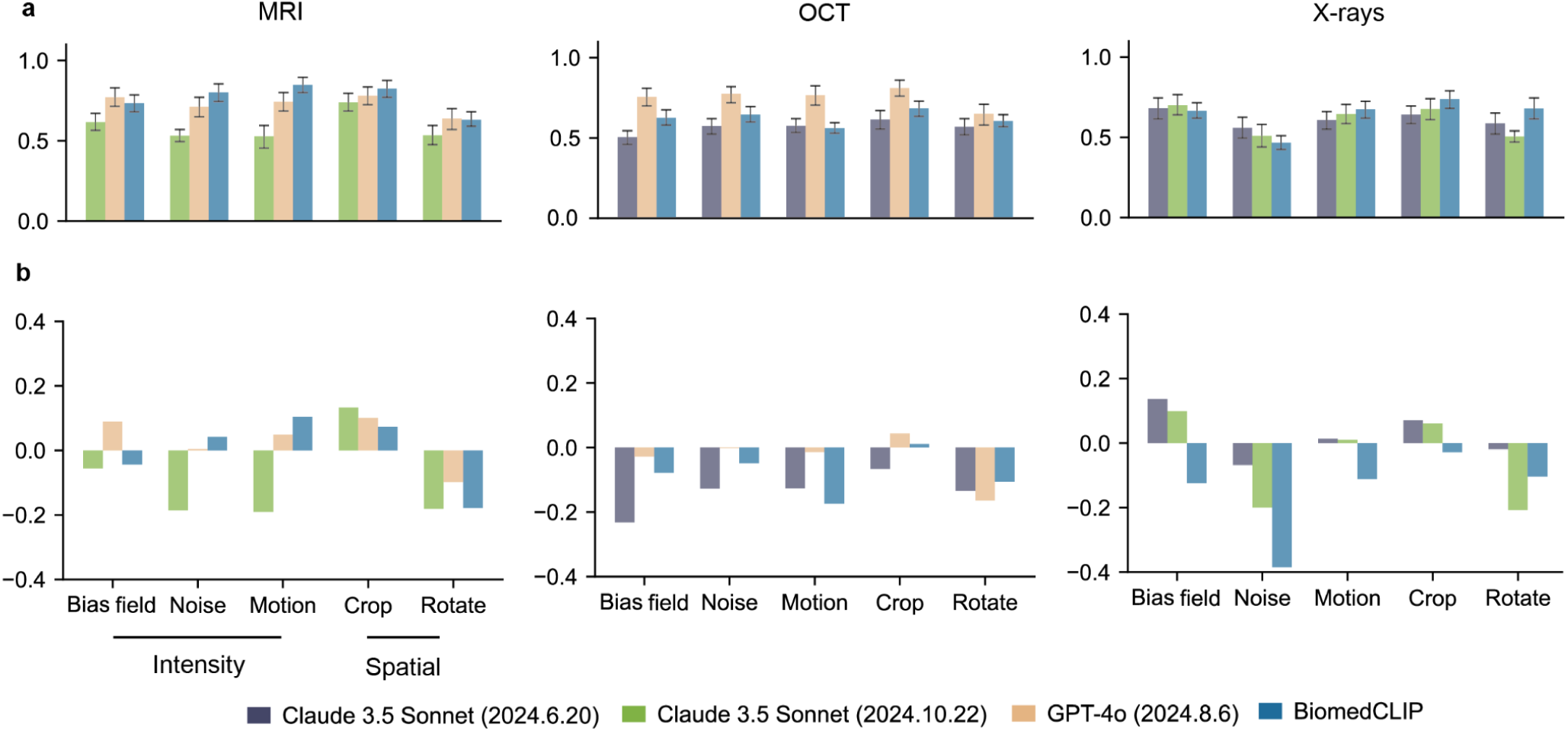
Vision-Language models’ (VLMs) accuracy in detecting disease on images containing weak artefacts using standard prompts. The top three models for each modality, when detecting disease from original unaltered images using standard prompts (Fig. 3 and Extended Data Figs. 4, 5), were included. Figure **a** shows the models’ accuracy (y-axis) after adding weak artefacts to the images. Full models performance through various prompt strategies shown in Extended Data Figs. 6∼8. Figure **b** shows a drop in model accuracy (y-axis) after the addition of weak artefacts. Model performance percentage change through various prompt strategies shown in Extended Data Figs. 9∼17. The full quantitative results and p-values for model performance at different scales of image artefacts are provided in Supplementary Table 8. For each disease detection task, we performed 1,000 iterations of stratified bootstrapping to calculate accuracy and sensitivity. Results show mean performance with 95% confidence intervals (error bars).

To better visualise the impact of weak artefacts, we plotted the performance percentage change for the top three models (as in Fig. 4a) relative to their original performance, as shown in Fig. 4b. We found that adding weak artefacts could, in some cases, increase model performance. In particular, random cropping usually improved model accuracy compared to other artefacts. For instance, BiomedCLIP’s macular disease detection accuracy increased by 1.1% (p<0.001) when random cropping was applied to OCT images, compared to a substantial drop of 17.4% (p<0.001) when random motion was introduced (Supplementary Table 9). Random noise led to more substantial decreases in sensitivity, particularly in MRI and OCT applications (Extended Data Figs. 9∼14). For the chest X-ray application, no specific artefact had a notably greater impact on model performance. However, artefacts generally caused the Claude series models to classify more cases as diseased, resulting in increased sensitivity but a slight drop in accuracy when using the standard prompt (Extended Data Figs. 15∼17).

### Strong artefacts detection rate

We assessed the VLMs’ ability to detect strong artefacts in severely distorted images (Fig. 5). We presented results for three models, as BiomedCLIP only outputs probabilities for each category, and Llama 3.2 11B classified most cases as diseased (Fig. 3 and Extended Data Figs. 4, 5). Claude 3.5 Sonnet (2024.6.20) achieved the highest strong artefacts detection rate in all medical fields using the standard prompt. For example, when analysing brain MRI slices with strong random motion, it achieved a detection rate of 0.825 (95% CI: 0.775 to 0.875), outperforming both its upgraded model (0.629; 95% CI: 0.565 to 0.695) and GPT-4o (0.230; 95% CI: 0.175 to 0.285) when using the standard prompt. Compared to the Claude series models, GPT-4o tended to refuse to answer rather than flagging poor image quality in MRI and X-ray applications, while few models refused to answer in OCT applications (Extended Data Fig. 18). Intensity artefacts were generally more easily detected by VLMs. For instance, Claude 3.5 Sonnet (2024.6.20) achieved a 0.945 (95% CI: 0.910 to 0.975) detection rate when analysing MRI slices with strong random noise, while showing no detecting capability 0.000 (95% CI: 0.000 to 0.000) for images with strong rotation.

**Fig. 5:**
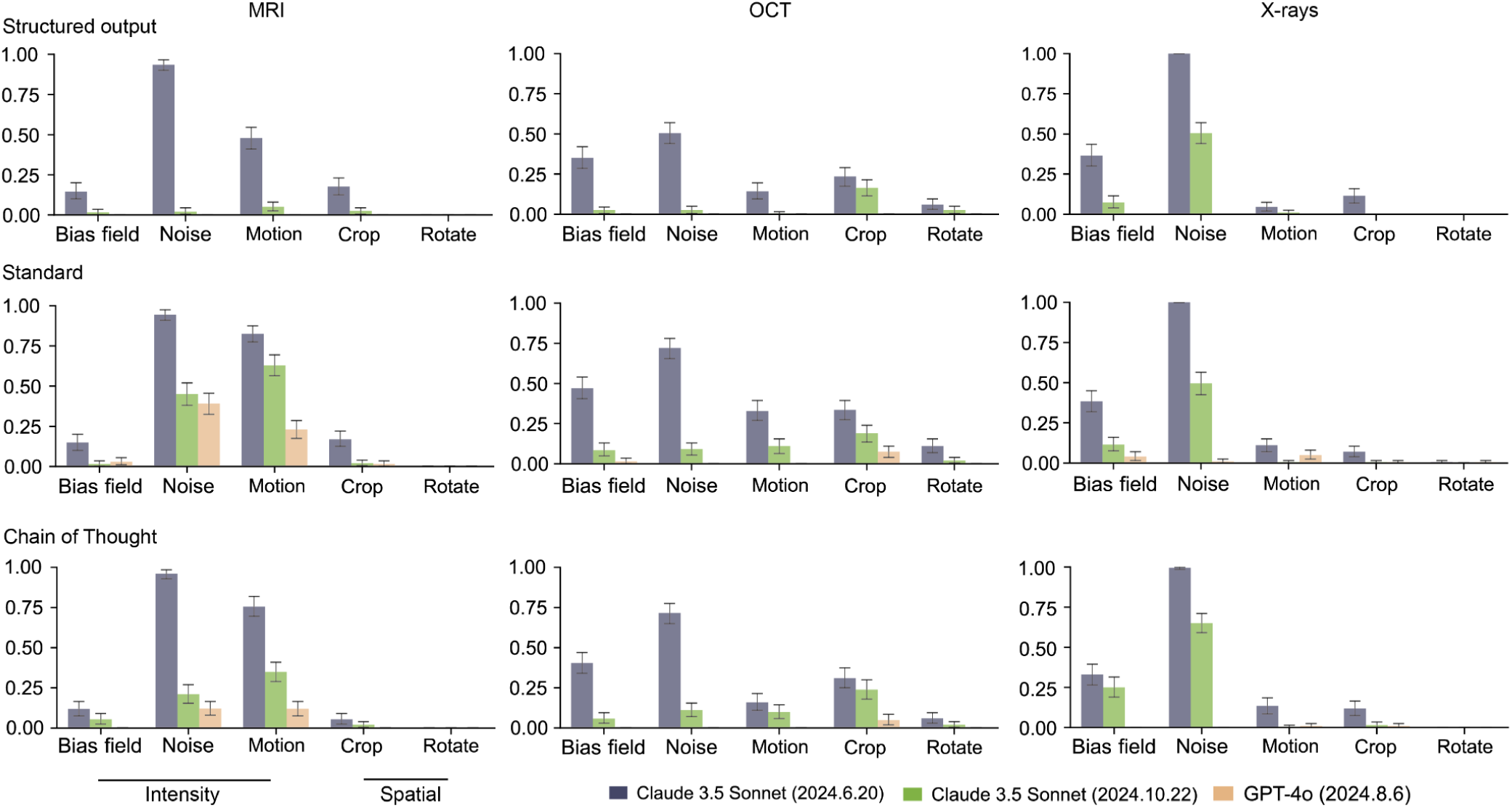
Vision-Language models’ (VLMs) detection rate for strong artefacts. The rate that VLMs flag poor image quality when analysing images distorted by strong artefacts is shown in the y-axis. We excluded Llama due to poor accuracy of diagnosing disease on original unaltered images (Fig. 3 and Extended Data Figs. 4, 5) and BiomedCLIP because it only outputs probability. Quantitative results are presented in Supplementary Table 10. Results show mean detection rates with 95% confidence intervals (error bars).

### Role of prompt engineering

We analysed the impact of prompt engineering to VLMs’ performance. When detecting original unaltered images, prompt engineering does not have an obvious impact on most models, except GPT-4o. GPT-4o’s accuracy and sensitivity decreased in the chest X-ray application when switching from a structured output to the Chain of Thought (CoT) prompt. Its sensitivity dropped from 0.750 (95% CI: 0.660 to 0.830) with the structured output prompt to 0.151 (95% CI: 0.090 to 0.220) with CoT (Extended Data Fig. 4). The impact of prompt engineering became more evident with weak artefacts, as the sensitivity drop caused by CoT in GPT-4o extended from chest X-rays to MRI and OCT applications, while other models were less affected. (Extended Data Figs. 6∼8). After adding stronger artefacts, standard prompts were better at encouraging models to flag poor quality images than other prompts. For instance, in MRI applications, Claude 3.5 Sonnet (2024.10.22) performed better with the standard prompt for detecting images with random noise, with rate of 0.629 (95% CI: 0.565 to 0.695), compared to 0.051 (95% CI: 0.025 to 0.080) with the structured output prompt (Fig. 5).

### Model performance on colour fundus photographs with real-world artefact

We assessed VLM robustness in detecting lesions from colour fundus photographs that contain real-world artefacts of varying scales, using the standard prompt (Extended Data Fig. 19). When detecting high quality images, GPT-4o had the highest accuracy 0.731 (95% CI: 0.660, 0.800) and highest specificity 0.960 (95% CI: 0.900, 1.000). While Llama 3.2 11B had the lowest accuracy 0.470 (95% CI: 0.430, 0.500), as it classified most cases as diabetic retinopathy, showing the highest sensitivity but lowest specificity. When detecting images with weak artefacts, most models had a drop in accuracy, excluding Llama 3.2 11B with a slight increase. Image artefacts had the largest impact on Claude 3.5 Sonnet (2024.6.20), showing the largest sensitivity increase 37.8% but specificity decrease 35.4%. When detecting ungradable images, Claude 3.5 Sonnet (2024.6.20) had the highest strong artefact detection rate 0.560 (95% CI: 0.460, 0.650). GPT-4o had the highest refuse to answer rate 0.259 (95% CI: 0.180, 0.350).

## Discussion

This study presents a comprehensive analysis of the robustness of popular VLMs in disease detection, particularly when handling medical images with various artefacts. The results reveal that most VLMs, particularly open-source general-purpose models, perform poorly on these disease detection tasks. The introduction of weak artefacts, particularly intensity artefacts, resulted in decreased performance across most VLMs. Furthermore, when strong artefacts were applied, few VLMs were able to recognize the degraded image quality. Comparable results were observed with real-world artefacts, underscoring that current VLMs lack sufficient robustness to handle images of suboptimal quality.

VLMs do not perform well on disease detection tasks, and perform even worse when artefacts are added to the original unaltered images. As robustness is a critical attribute of AI models, it is essential to emphasize this aspect from the early stages, even when models still struggle to detect diseases in medical images of acceptable quality. Recent works have tested VLMs’ performance in disease detection, and indicated that GPT-4o usually achieves the optimal performance compared to other models^25,27,28,35^. In this study, we found that Llama 3.2 11B had the lowest disease detection accuracy across all medical fields. GPT-4o showed good sensitivity (Extended Data Fig. 4), similar to the observation by Johri et al^36^. However, we found that GPT-4o’s accuracy was not always the highest and fell behind BiomedCLIP in the chest X-ray applications (Fig. 3). This could underscore the importance of pre-training in the medical field before deploying VLMs for disease detection tasks. Few studies have evaluated VLM performance on medical images with added artefacts. Most recent work focuses on robustness to adversarial attacks^37–39^, while only a limited number examine artefacts—mainly random noise—highlighting VLMs’ poor robustness^27^. This mirrors our findings. More specifically, VLMs show poorer robustness against intensity artefacts, particularly random noise. This may be because spatial artefacts preserve more features compared to intensity artefacts (Extended Data Figs. 1∼3). Interestingly, VLM performance usually improves with random cropping. This enhancement might be attributed to either the cropping highlighting lesions by removing normal tissue, or these models may be better at processing lower resolution images (Extended Data Fig. 1d). Additionally, despite image rotation only altering its orientation, this leads to performance degradation across all applications in our study, probably due to training on single-orientation images. When detecting colour fundus photographs with weak real-world artefacts, we observed that the accuracy of some models (e.g., Claude 3.5 Sonnet 2024.6.20) decreased but exhibited increased sensitivity compared to their performance on high-quality images. This is likely because these models misinterpret artefacts as lesions.

Few models are capable of flagging strong image artefacts in disease detection tasks. Recent studies evaluating VLMs often use public datasets with acceptable image quality, overlooking their ability to identify poor-quality images before detecting lesions^13,15,16,40,41^. In this study, we find that VLMs are better at flagging intensity artefacts. This is probably because intensity artefacts largely change the intensity distribution compared to spatial artefacts (e.g., rotation). In the intensity artefacts, we find that VLMs are better at identifying severely distorted images caused by random noise (Fig. 5), likely due to their ability to recognize Gaussian distributions in image intensity distribution patterns. In contrast, other intensity artefacts such as the random bias field, which do not follow Gaussian distributions, present greater detection challenges for VLMs and require enhanced processing methods.

VLMs show inconsistent disease detection performance across medical fields. VLMs demonstrate higher accuracy in OCT applications compared to MRI, but exhibit lower sensitivity. This discrepancy may be attributed to the larger lesion sizes in brain MRI, which make abnormal regions more detectable. In contrast, lesions in OCT are typically smaller and less distinguishable from normal tissue, resulting in reduced sensitivity. When detecting lesions on images with weak artefacts, VLMs show reduced sensitivity in MRI and OCT applications but increased sensitivity in X-rays. This paradox may stem from opacity being a key marker for pulmonary lesions—since healthy lungs appear with low opacity, artefacts that increase opacity may bias VLMs toward diseased predictions (Extended Data Fig. 3), raising sensitivity. Compared to chest X-ray applications, VLMs detect strong artefacts more effectively in MRI. One possible explanation lies in the differing image complexity and intensity distributions. Chest X-rays typically exhibit a simpler intensity profile—primarily dark lung regions against bright anatomical backgrounds—which can make artefacts such as motion blur less distinguishable from normal anatomical variation. In contrast, brain MRI presents more complex structural details and greater variability, where artefacts that disrupt structural boundaries become more visually salient.

The VLMs performance improvement through Chain of Thought (CoT) is hardly seen in disease detection tasks. Previous works report that CoT prompting can improve the VLMs’ performance on general tasks, particular tasks relying on logical thinking^42–46^. Whether this advantage extends to the medical domain, specifically in disease detection, has not been fully tested. In this study, some VLMs’ performance deteriorated with CoT prompting compared to structured output prompt, particularly GPT-4o (Fig. 3 and Extended Data Figs. 4, 5). The decline in CoT is attributed to categorizing both refusing to answer and flagging poor image quality responses as incorrect answers, specifically as GPT-4o tends to refuse to answer more frequently using CoT prompt (Extended Data Fig. 18). While CoT does not increase the rate of refuse to answer for most VLMs when analysing original unaltered images, their performance improvement is still marginal. When VLMs detect diseases through standard and CoT prompts, we usually find that these models have enough medical knowledge but still make mistakes. This may be because they are hard to capture pathological features from medical images at latent space, although they can analyse the intensity distribution of them.

A comprehensive evaluation of VLMs’ disease detection performance—specifically robustness—is essential for advancing their development and explainability. Ideally, medical VLMs should emulate clinicians by maintaining stable diagnostic accuracy when images are slightly distorted and recommending re-examinations when the images are ungradable. However, current models fall short in both aspects, with the assessment of image quality prior to lesion detection remaining largely neglected. Future VLMs should integrate a quality assessment module before diagnosis. Furthermore, our results may also inspire researchers to explore the reasons behind VLMs’ suboptimal performance. For instance, future work could visualise the attention map from vision encoders to see if meaningful pathological features are captured at different medical images, particularly for those with small lesion size (retinal OCT images in our case). Although the advantages of CoT reasoning may not be immediately observable in current applications, prompt engineering holds promise for improving disease detection performance. Ferber et al. showed that in-context learning improves VLMs’ classification of cancer pathology images^47^. More broadly, systematic exploration of advanced prompting strategies may lead to further performance gains.

By building the VLMs’ robustness evaluation framework, we hope to accelerate the progress to better understanding and developing robust medical VLMs. Several limitations of this study warrant consideration. First, we introduced five categories of image artefacts commonly encountered in clinical scenarios. However, these may not fully capture the complexity of artefacts found in real-world medical imaging, where artefacts often appear in combination rather than isolation. It is worth exploring model robustness to combinations of image artefacts. Second, the intensity levels for weak and strong image artefacts were preset hyperparameters. Future research should provide a quantified threshold for weak and strong artefacts (e.g., quantifying the difference between images with weak artefacts and their original unaltered counterparts), therefore a more fine-grained scale of image artefacts will be explored. Third, disease detection tasks are generally simplified into a two-category format (normal vs. diseased), which is relevant to the poor performance of VLMs, even in these simplified cases (Fig. 3). The future work will further refine the VLM robustness evaluation framework by simulating more realistic image artefacts, exploring fine-grained artefact scales, and incorporating multi-class disease classification tasks.

We build a benchmarking framework to evaluate VLMs’ robustness to image artefacts in three disease detection tasks across multiple medical fields. By pinpointing vulnerabilities tied to specific medical fields and artefact types, this work provides insights for developing robust medical VLMs capable of functioning reliably in real-world clinical settings.

## Method

### Benchmark Dataset

The Brain Tumour MRI Dataset^48^ includes images of three tumour types—gliomas, meningiomas, and pituitary tumours—along with healthy scans. It combines three sources: Figshare^49^, the SARTAJ dataset^50^, and Br35H^51^. Figshare images were acquired post-Gd-DTPA injection at Nanfang Hospital and Tianjin Medical University Hospital (2005.9∼2010.10); the other sources lack detailed provenance. Patient age and sex are not provided. Each MRI scan yields multiple slices, labelled by experienced radiologists. For this study, we randomly selected 100 images each from the three tumour types to form the tumour class, and 100 healthy images to form the normal class.

The Retinal OCT Images dataset^52^ includes three macular diseases—choroidal neovascularization (CNV), diabetic macular oedema (DME), and drusen—alongside normal scans. Images were collected from retrospective adult cohorts at five global institutions between July 1, 2013, and March 1, 2017. Patient age and sex are not provided. Multiple slices per scan were labelled through a tiered grading process involving medical students, ophthalmologists, and senior retinal specialists with over 20 years of experience. For this study, we randomly selected 100 images each from the three disease types to form the macular diseases class, and 100 images from class without lesions to build the normal class.

The COVID-19 Detection X-ray Dataset^53^ includes three lung diseases—bacterial pneumonia, COVID-19, and viral pneumonia—alongside normal chest X-rays. COVID-19 images were sourced from public datasets^54^, while pneumonia and normal scans were from paediatric patients (ages 1∼5) at Guangzhou Women and Children’s Medical Centre^55^. Patient age, sex, and number of scans per patient are not specified. Diagnoses were reviewed by two expert physicians. For this study, we randomly selected 100 images each from the three disease classes to form the lung diseases class, and 100 images from class without lesions to build the normal class.

For each dataset, we introduced five types of artefacts—each at weak and strong levels—to the original images (Supplementary Table 2). Weak artefact datasets contain ∼30% ungradable images, with the remaining 70% partially obscured but still interpretable. Strong artefact datasets include ∼90% ungradable images. Intensity artefacts (random noise, motion, bias fields) simulate issues from hardware noise and operational factors, applied using the TorchIO library^56^. Spatial artefacts (rotation, cropping) mimic misalignment and limited field of view during acquisition (Extended Data Figs. 1∼3). Details are in Supplementary Table 3. Questions were generated from ground truth labels to convert the data into VQA tasks.

To better demonstrate VLMs’ robustness, we included images with real-world artefacts from the Diabetic Retinopathy (DDR) dataset in the color fundus modality, which contains ungradable images^57^. Collected from 147 hospitals across 23 provinces in China, the dataset lacks details on patient age, sex, and scan count. Images were annotated by ophthalmologists and classified into six DR severity levels: none (‘0’), mild (‘1’), moderate (‘2’), severe (‘3’), proliferative (‘4’), and ungradable (‘5’). Due to the subtlety of mild DR, we grouped mild DR and non-DR (‘0’∼‘1’) as normal, ‘2’∼‘4’ as diseased, and ‘5’ as strongly artefactual. We selected 50 high-quality images from both the normal and diseased classes, respectively. Therefore, we selected 50 low-quality images from each of the normal and diseased classes to construct the weak artefact dataset, and 100 images from class ‘5’ to represent the strong artefact dataset.

### Metrics for model robustness

We manually categorized VLM responses into four classes: ‘0’ for those indicating normal findings without lesions, ‘1’ for responses indicating image abnormalities, ‘2’ for any cases where VLMs flag poor image quality, and ‘3’ for instances where VLMs refused to answer due to ethical concerns or suggested consulting a specialist. All results and manual annotations are provided in Supplementary Table 4. We used three metrics to evaluate VLM robustness.

#### Model performance

The accuracy and sensitivity of VLMs in disease detection. Some responses from VLM did not provide a specific answer between disease and normal (with answers ‘2’ and ‘3’). We processed them as incorrect answers in calculating accuracy and sensitivity.

#### Performance percentage change

The performance difference between detecting diseases on images with weak artefacts and on original unaltered images, divided by original performance. For example, if GPT-4o’s accuracy in detecting brain tumours on original MRI is 0.7, and after adding weak random noise, the accuracy declines to 0.5, the performance percentage change is -28.6%.

#### Strong artefacts detection rate

The proportion of responses that flagged poor image quality in images with strong artefacts (with answer ‘2’), compared to the total number of responses.

## Models evaluated

This included three proprietary models—GPT-4o (2024.8.6)^2^, Claude 3.5 Sonnet (2024.6.20)^3^, and the upgraded version Claude 3.5 Sonnet (2024.10.22). Claude 3.5 Sonnet was upgraded during the experiment, we hence tested the performance of both versions. Additionally, we tested two open-source models, Llama 3.2 with 11 billion parameters^58^, and BiomedCLIP^7^, an open-source VLM specifically designed for medical classification tasks, which outputs probabilities for each category. To maintain reproducibility, avoiding different responses for the same disease detection task, we set the temperature parameter as 0 for GPT-4o and both versions of Claude 3.5 Sonnet. For Llama 3.2, the temperature was configured to 0.01 to avoid the error that occurs when it is set to 0. BiomedCLIP does not define temperature parameters.

We tested three classical prompt strategies given that prompt substantially impacts VLMs’ performance: (1) structured output prompt, restricting the model to output only the categorical answer (e.g., “diseased” or “normal”); (2) standard prompt, a default prompt style allowing open answering without special restrictions; and (3) Chain of Thought (CoT) prompt^42^, including instructions such as “Describe your reasoning step by step” to encourage detailed, sequential reasoning. The details of these prompt strategies are put in the Supplementary Table 5.

## Statistical analysis

We employed stratified bootstrapping with 1,000 iterations to maintain class distributions. We calculate the mean and standard deviation of the performance over the 1000 iterations and calculate the standard error by (standard deviation/ 5). We obtain the 95% CI by means of 1.96 × standard error. To assess the statistical significance of performance changes due to adding artefacts, we conducted the two-tailed t-test to calculate p-values and put the result in Supplementary Table 9.

## Data availability

The benchmark dataset, which includes three imaging modalities with introduced artefacts, is derived from publicly available sources and can be accessed via the following links: MRI (https://www.kaggle.com/datasets/masoudnickparvar/brain-tumor-mri-dataset), OCT (https://www.kaggle.com/datasets/paultimothymooney/kermany2018), and X-ray (https://www.kaggle.com/datasets/darshan1504/covid19-detection-xray-dataset). We have reorganized, randomly sampled, and added artefacts to create this benchmark, which is available at: https://github.com/ziijiecheng/VLM_robustness. Additionally, the colour fundus dataset, containing real-world artefacts, is available at: https://github.com/nkicsl/DDR-dataset. We have reorganized and sampled it according to image distortion levels, and it can be found alongside the benchmark dataset.

## Code availability

The code used to introduce artefacts and evaluate the performance of VLMs from Z.C. is available at: https://github.com/ziijiecheng/VLM_robustness. The code for introducing image artefacts uses TorchIO as the reference: https://github.com/TorchIO-project/torchio. The reference code for VLM evaluation is available for the following models: GPT-4o (https://platform.openai.com/docs/quickstart), Claude 3.5 Sonnet (https://github.com/anthropics/anthropic-cookbook), BiomedCLIP (https://huggingface.co/microsoft/BiomedCLIP-PubMedBERT_256-vit_base_patch16_224), and Llama 3.2 (https://huggingface.co/meta-llama/Llama-3.2-11B-Vision-Instruct). Results were further analyzed and visualized using Python v.3.9.18, NumPy v.1.26.2, Matplotlib v.3.8.0, SciPy v.1.11.3, and pandas v.1.5.0.

## Acknowledgement

P.A.K. is supported by a Moorfields Eye Charity Career Development Award (grant no. R190028A) and a UK Research & Innovation Future Leaders Fellowship (grant no. MR/T019050/1). For the purpose of open access, the author has applied a Creative Commons Attribution (CC BY) licence to any Author Accepted Manuscript version arising.

## Author contributions

Z.C., B.L., L.J., T.H., C.Y.C. and Y.Z. contributed to the conception and design of the work. Z.C. and Y.Z. contributed to the data organization, technical implementation and analysis framework. All authors contributed to the drafting and revising of the manuscript.

## Supplementary Material

Supplementary Table 1 False positive/negative examples

Supplementary Table 2 Benchmark summary

Supplementary Table 3 Image artefacts settings

Supplementary Table 4 All VLMs’ responses

Supplementary Table 5 Prompts’ detail

Supplementary Table 6 Quantitative results of VLMs’ original performance

Supplementary Table 7 Quantitative results of VLMs’ performance after adding weak artefacts

Supplementary Table 8 Quantitative results of VLMs’ performance percentage change after adding weak artefacts

Supplementary Table 9 p-values for model performance at different scales of image artefacts

Supplementary Table 10 Quantitative results of VLMs’ strong artefacts detection and refuse to answer rate

Supplementary Table 11 Quantitative results of VLMs’ robustness in color fundus photographs with real world artefact

**Extended Data Fig. 1:**
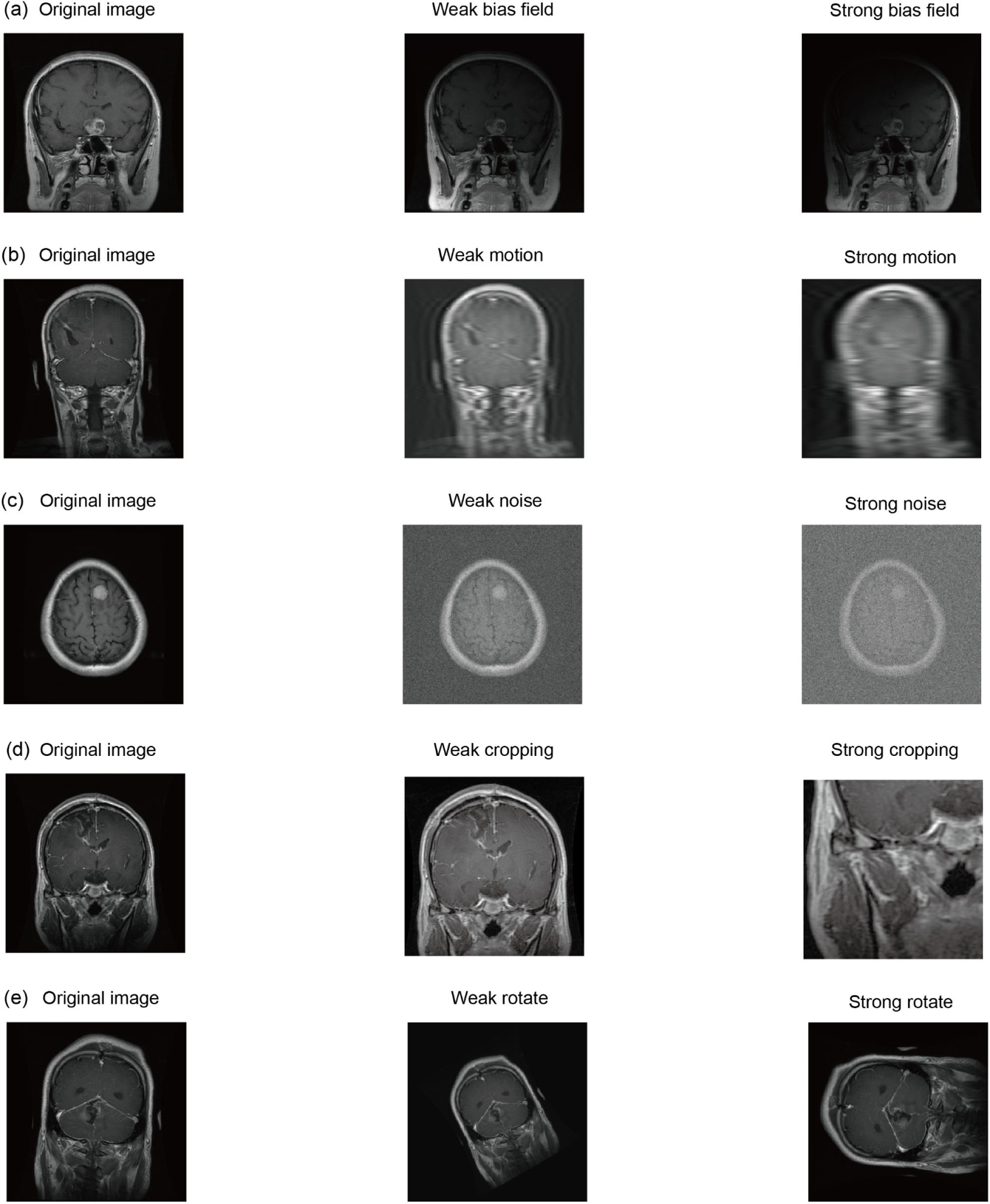
Examples of brain MRI slices with weak and strong artefacts.

**Extended Data Fig. 2:**
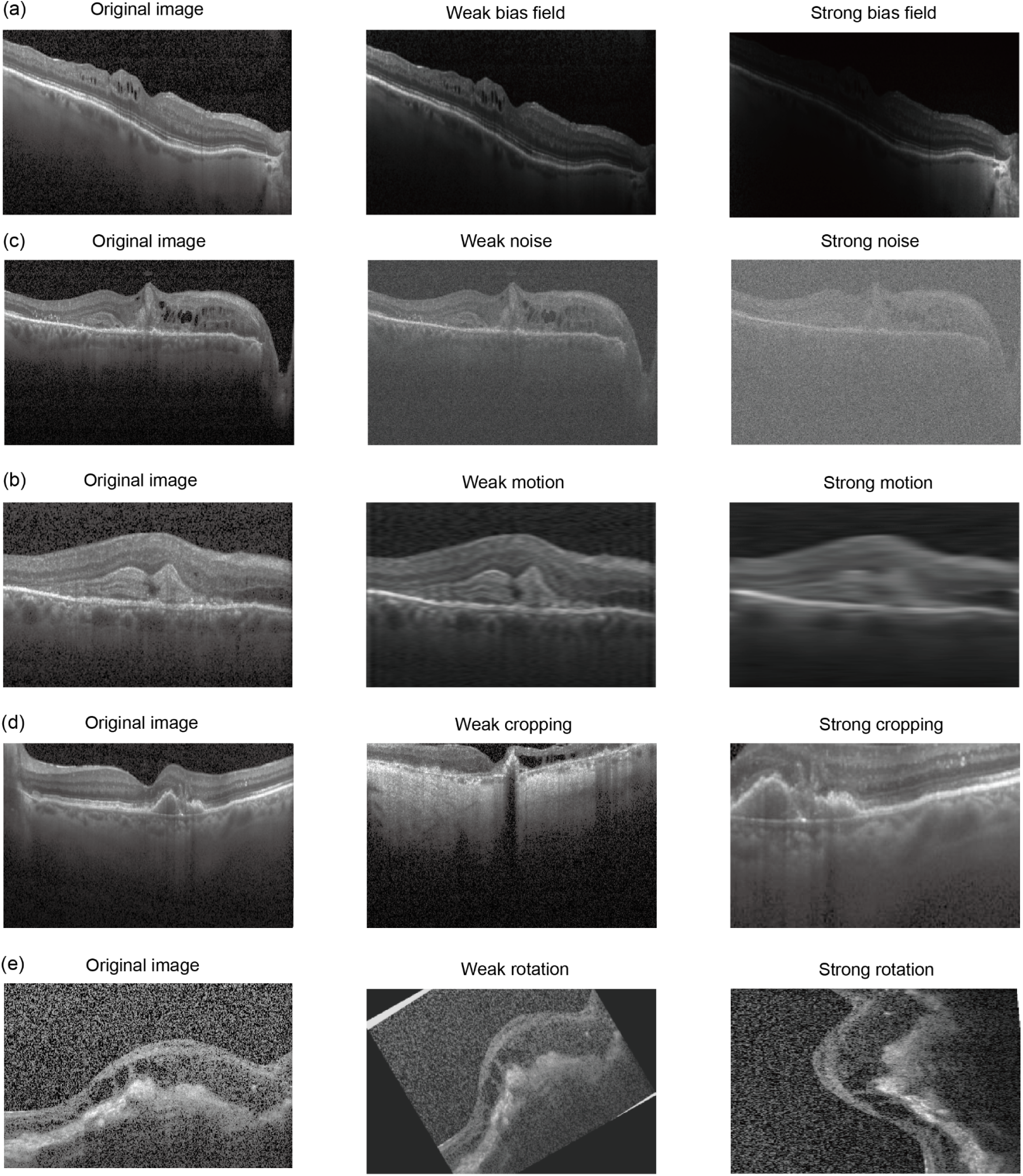
Examples of OCT slices with weak and strong artefacts.

**Extended Data Fig. 3:**
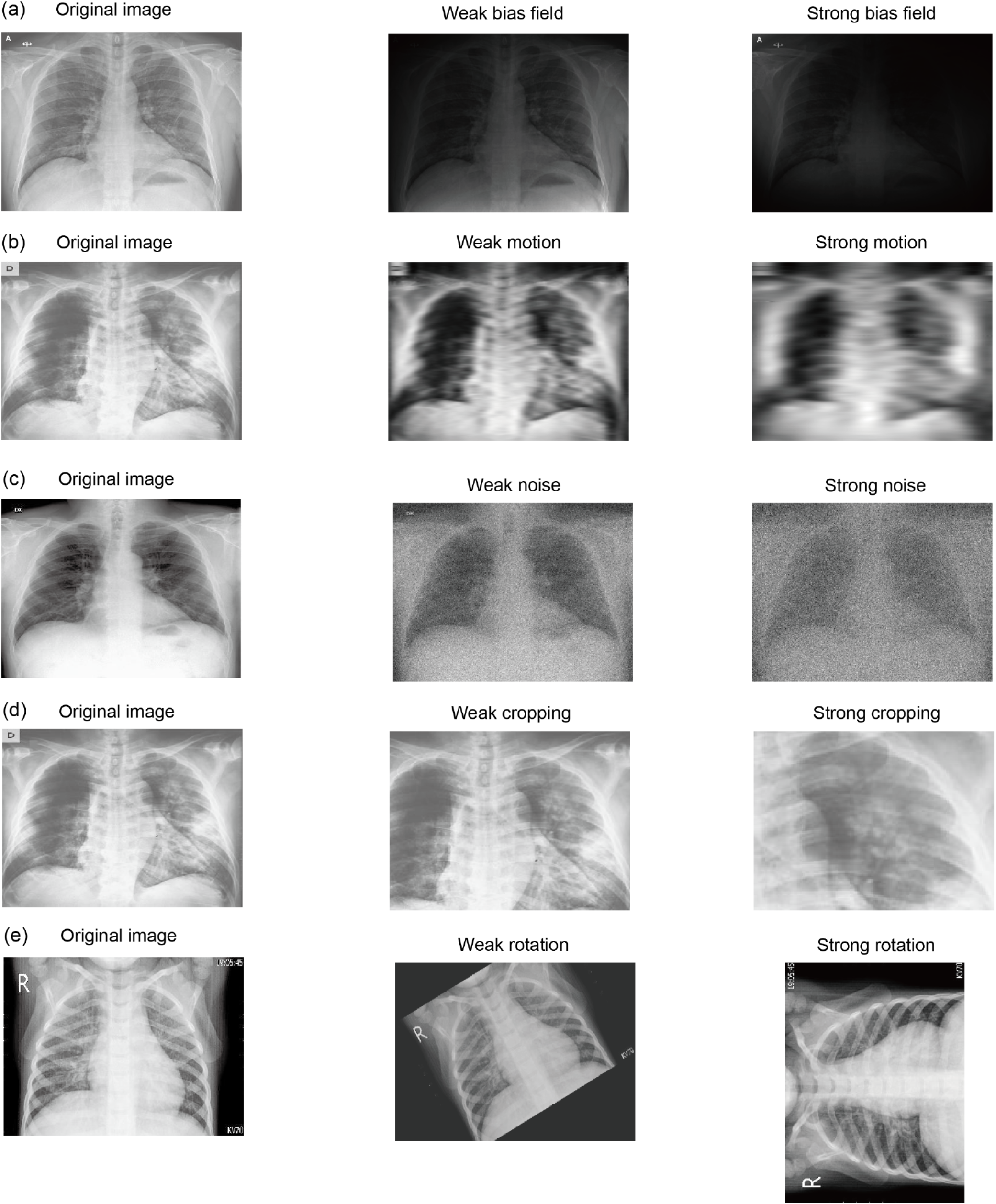
Examples of chest X-ray with weak and strong scales artefacts.

**Extended Data Fig. 4:**
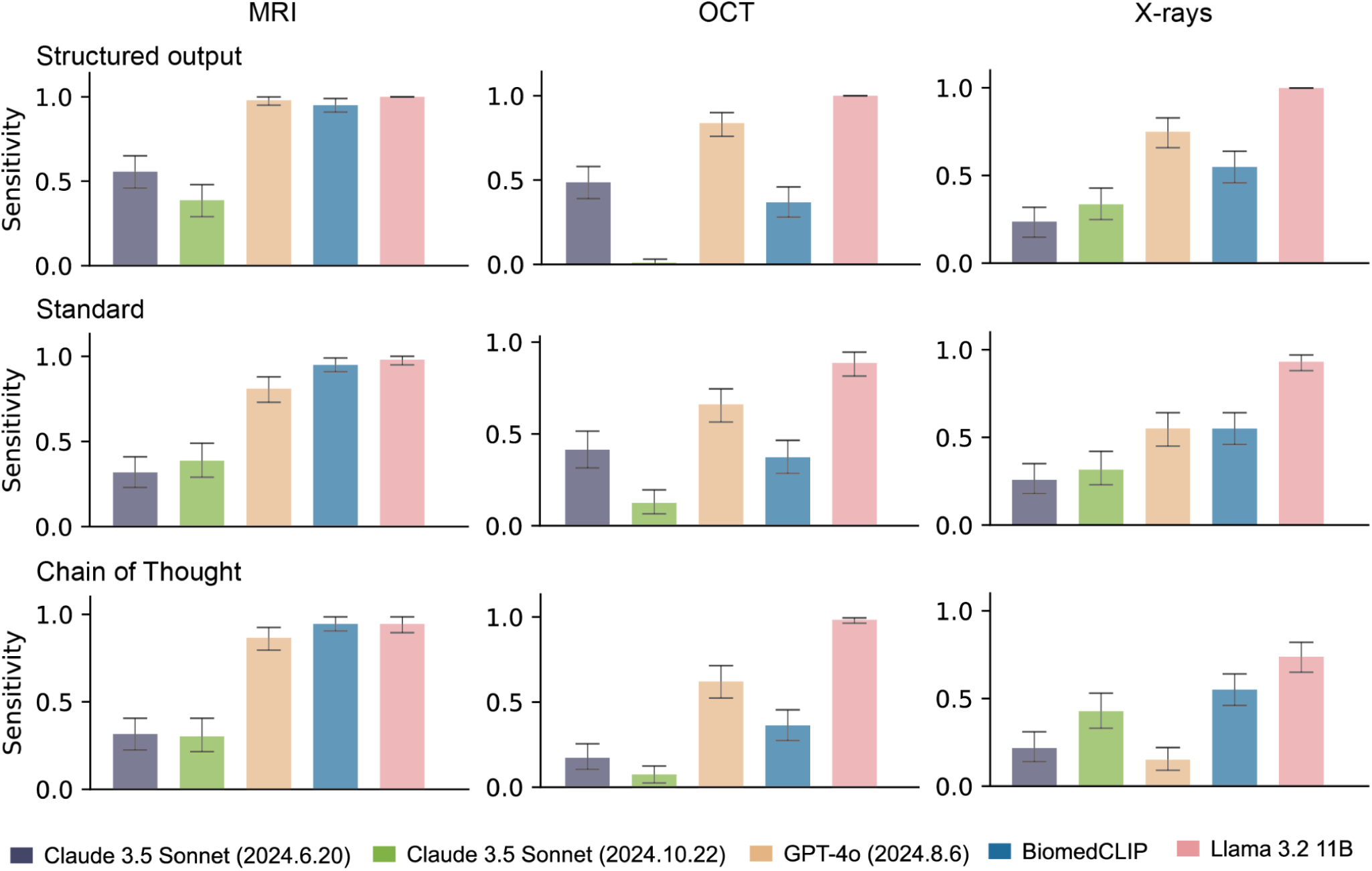
The sensitivity of Vision-Language models (VLMs) in detecting disease from original unaltered images. BiomedCLIP requires only task categories and outputs the probability for each category, but we include it across all prompt strategies for comparison. Quantitative **results** are detailed in Supplementary Table 6. For each task, sensitivity measurements were derived from 1000 stratified bootstrap samples. Results show mean sensitivity with 95% confidence intervals (error bars).

**Extended Data Fig. 5:**
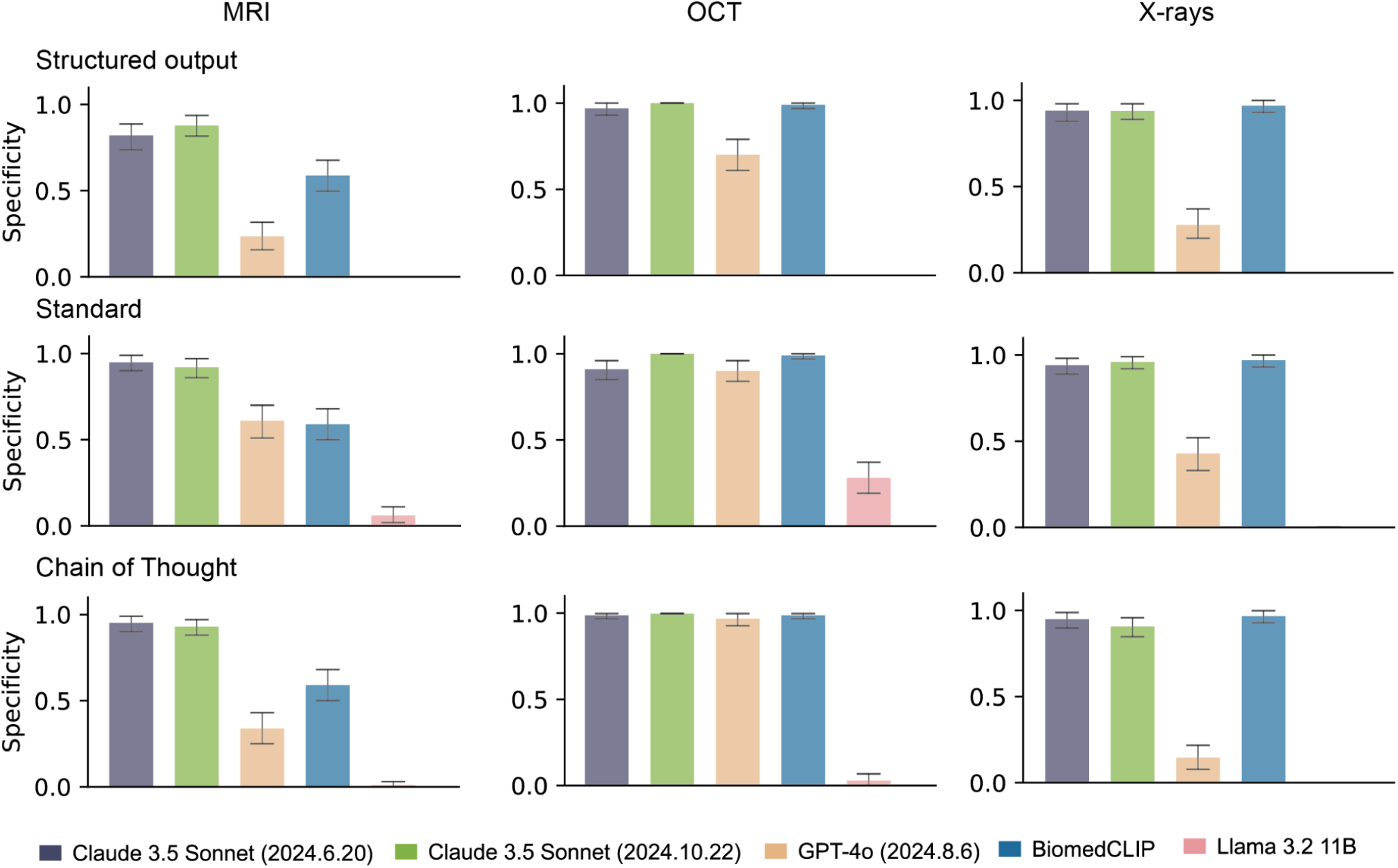
The specificity of Vision-Language models (VLMs) in detecting disease from original unaltered images. BiomedCLIP requires only task categories and outputs the probability for each category, but we include it across all prompt strategies for comparison. Quantitative **results** are detailed in Supplementary Table 6. For each task, specificity measurements were derived from 1000 stratified bootstrap samples. Results show mean specificity with 95% confidence intervals (error bars).

**Extended Data Fig. 6:**
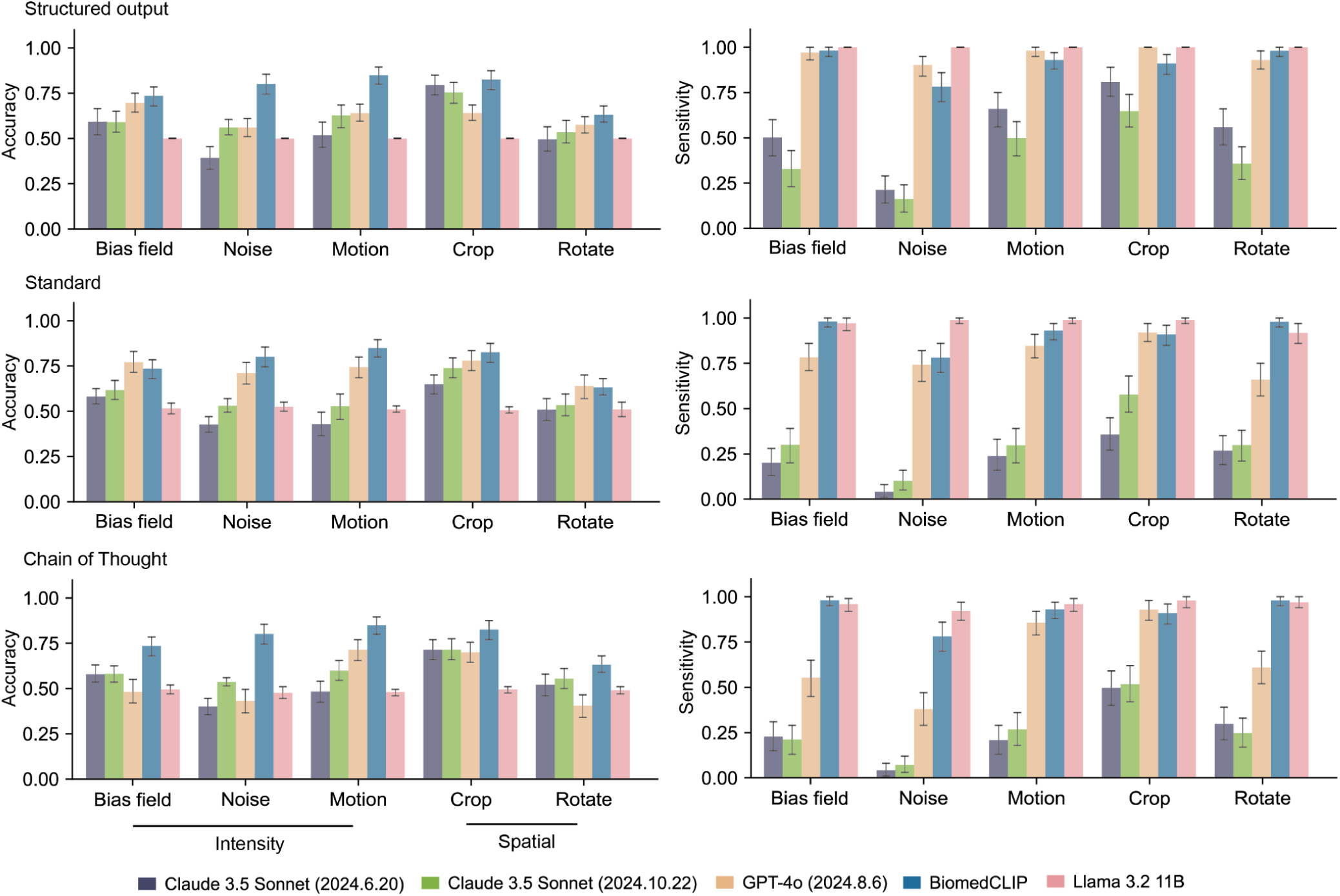
The model performance of all Vision-Language models (VLMs) in MRI applications after adding weak artefacts to the original unaltered images. Complete quantitative results for VLMs’ accuracy, sensitivity in brain tumour detection are available in Supplementary Table 7. For **each** disease detection task, we performed 1,000 iterations of stratified bootstrapping to calculate accuracy and sensitivity. Results show mean performance with 95% confidence intervals (error bars).

**Extended Data Fig. 7:**
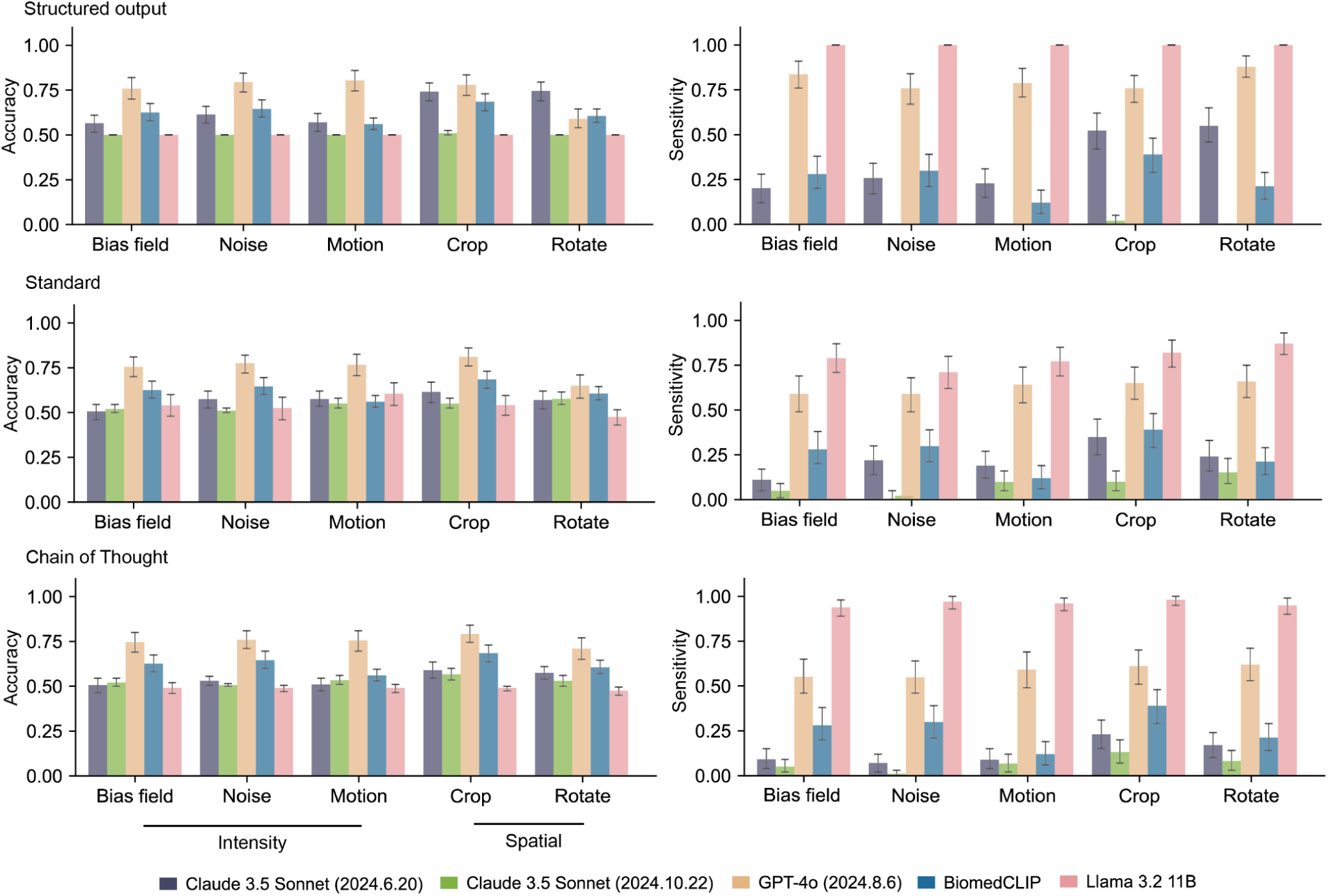
The model performance of all Vision-Language models (VLMs) in OCT applications after adding weak artefacts to the original unaltered images. Complete quantitative results for VLMs’ accuracy and sensitivity in macular disease detection are available in Supplementary Table 7. For each task, we performed 1,000 iterations of stratified bootstrapping to calculate accuracy and sensitivity. Results show mean performance with 95% confidence intervals (error bars).

**Extended Data Fig. 8:**
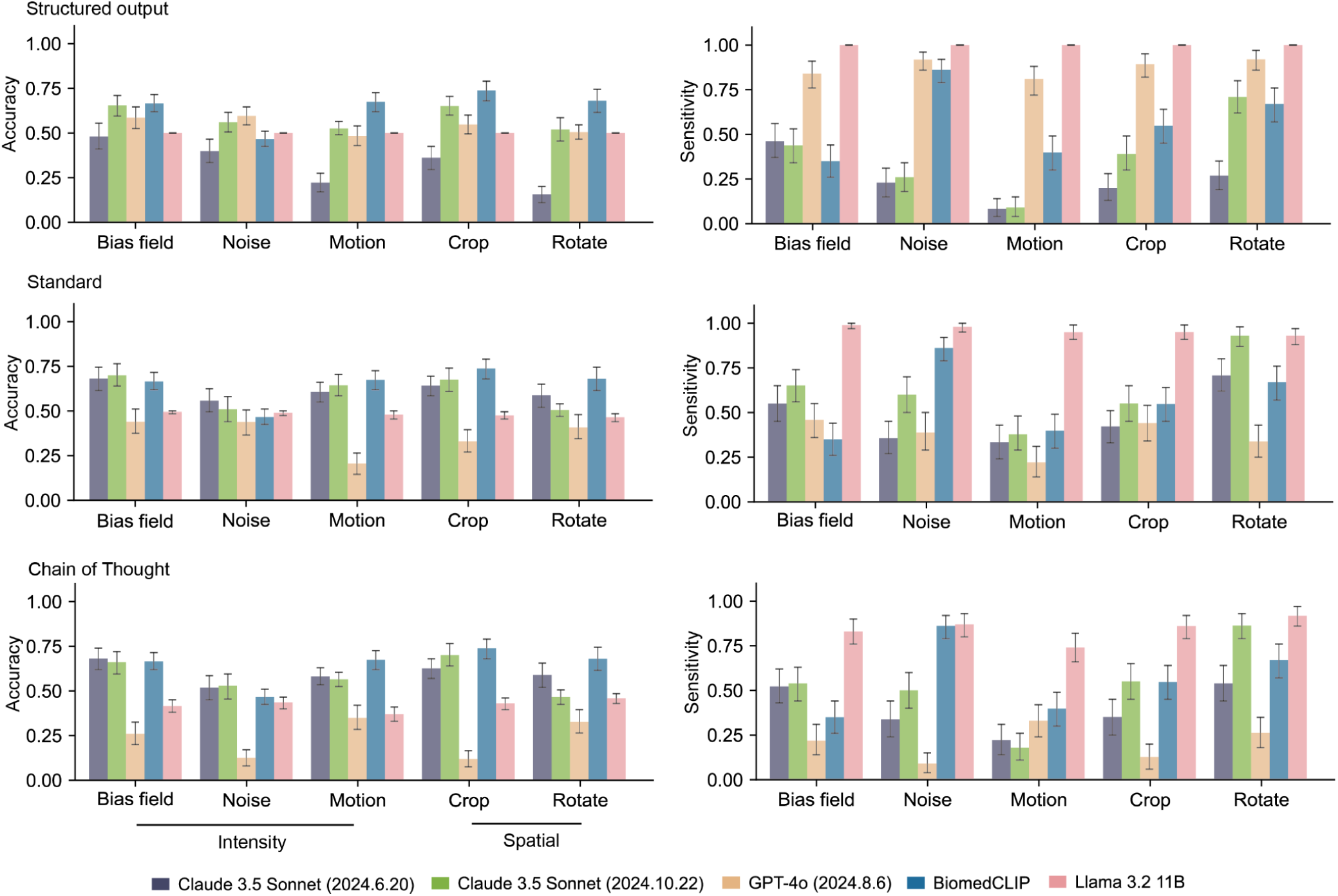
The model performance of all Vision-Language models (VLMs) in X-ray applications after adding weak artefacts to the original unaltered images. Complete quantitative results for VLMs’ accuracy, sensitivity in Covid/pneumonia detection are available in Supplementary Table 7. For each task, we performed 1,000 iterations of stratified bootstrapping to calculate accuracy and sensitivity. Results show mean performance with 95% confidence intervals (error bars).

**Extended Data Fig. 9:**
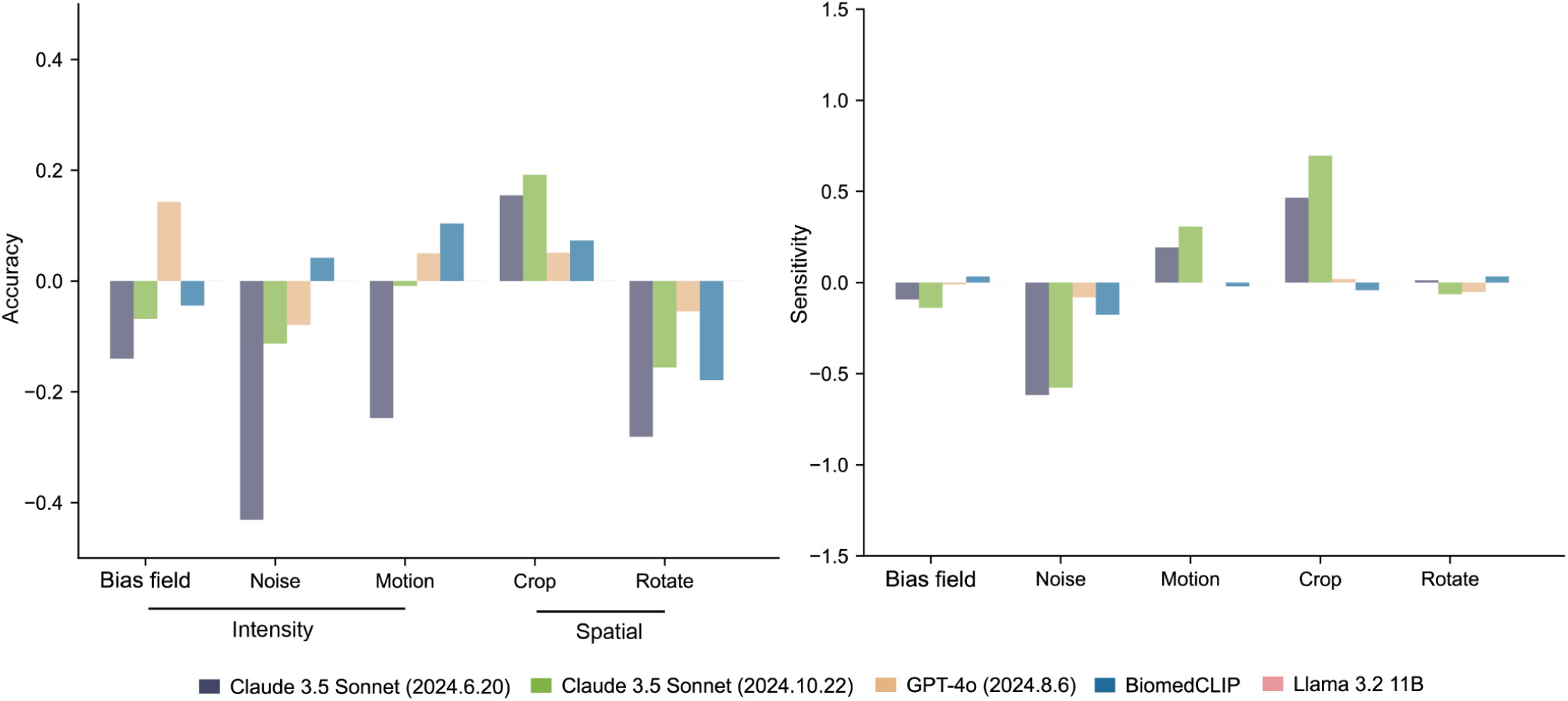
The deterioration in performance for all Vision-Language models (VLMs) after adding weak artefacts to original unaltered images in MRI applications using the structured output prompt. Performance percentage change due to adding weak artefacts to original unaltered images are represented in the y-axis. Complete quantitative data is available in Supplementary Table 8. For each task, we calculated performance percentage change through 1,000 iterations of stratified bootstrapping, with bar height representing mean value.

**Extended Data Fig. 10:**
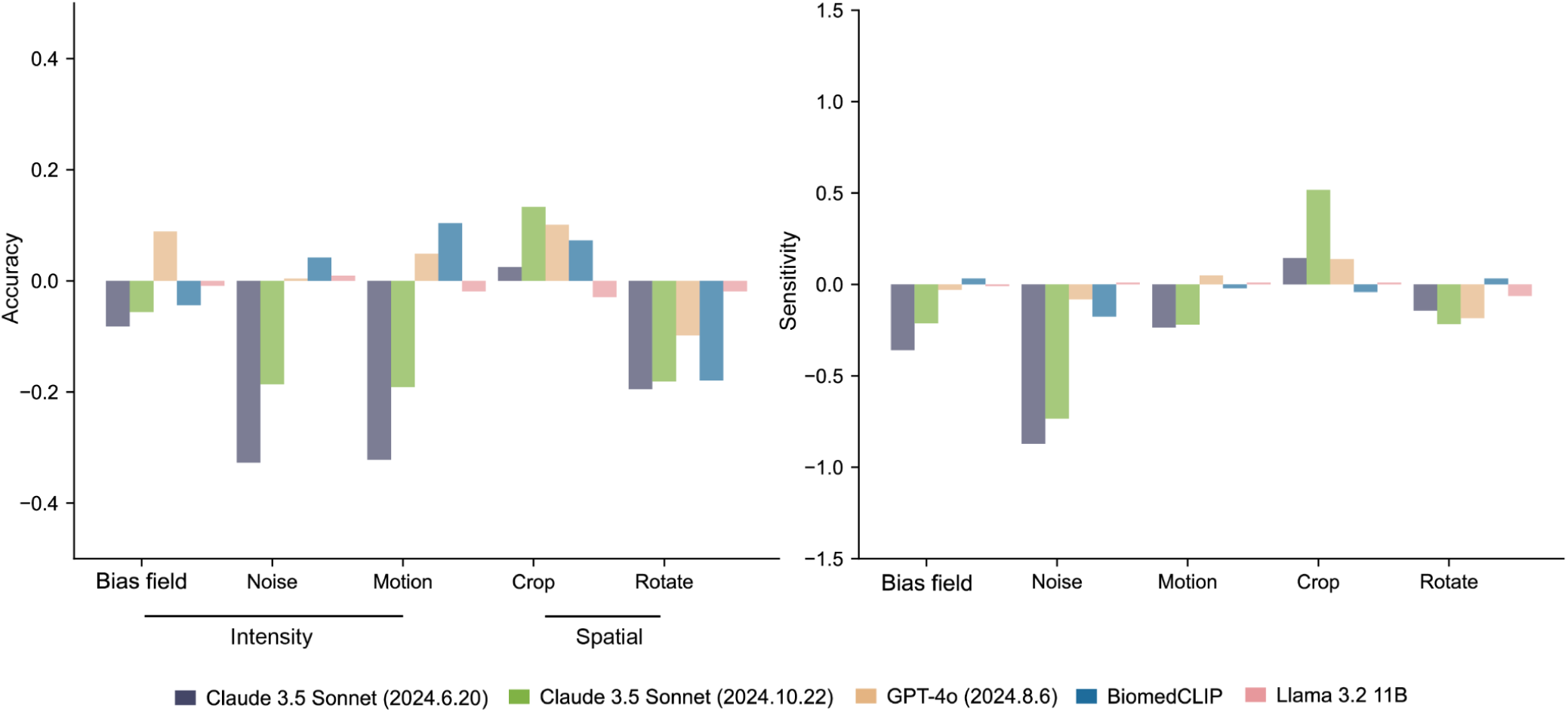
The performance percentage change of all Vision-Language models (VLMs) after adding weak artefacts to original MRI images using standard prompts. Performance percentage change due to adding weak artefacts to original unaltered images are represented as y-axis. Complete quantitative data is available in Supplementary Table 8. For each task, we calculated performance percentage change through 1,000 iterations of stratified bootstrapping, with bar height representing mean value.

**Extended Data Fig. 11:**
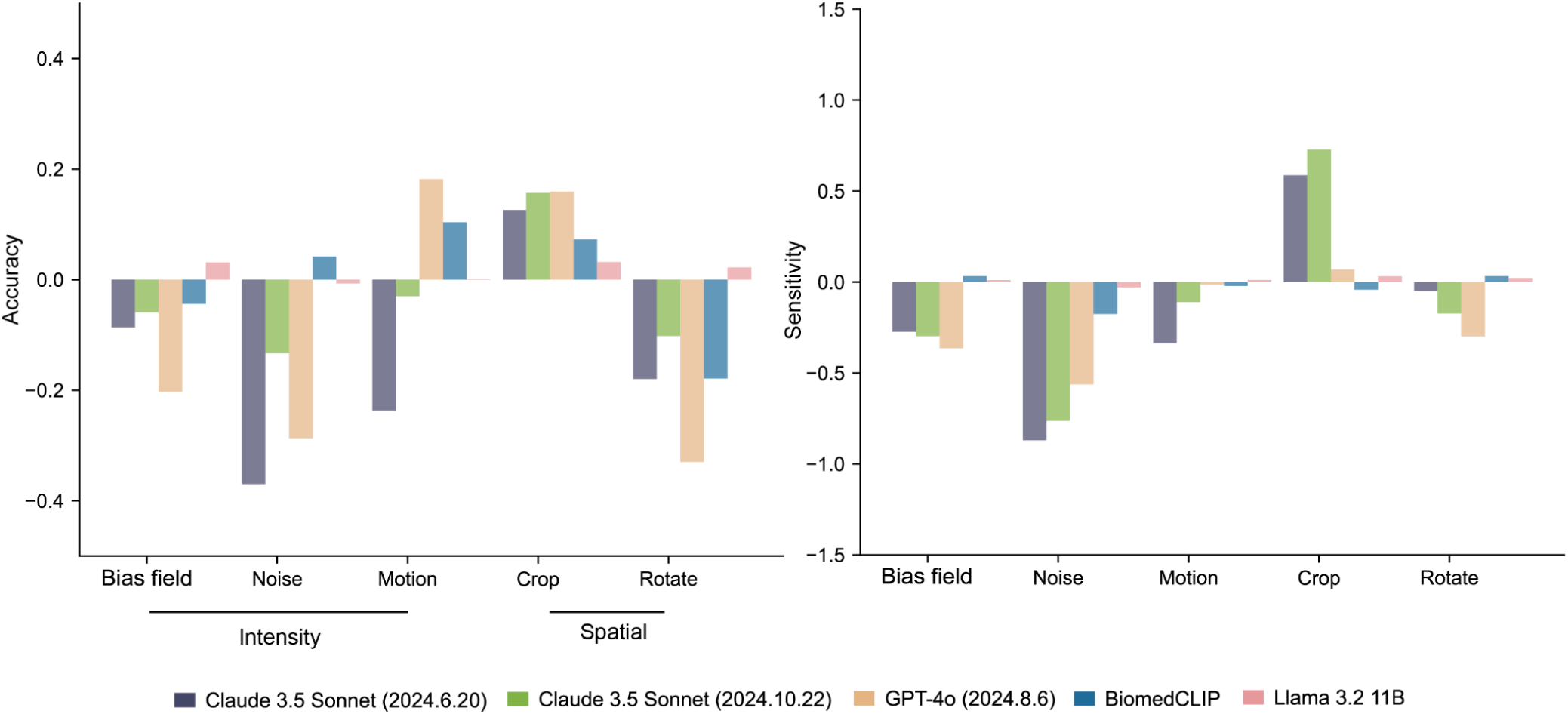
The performance percentage change of all Vision-Language models (VLMs) after adding weak artefacts to original MRI images using Chain of Thought prompts. Performance percentage change due to adding weak artefacts to original unaltered images are represented as y-axis. Complete quantitative data is available in Supplementary Table 8. For each task, we calculated performance percentage change through 1,000 iterations of stratified bootstrapping, with bar height representing mean value.

**Extended Data Fig. 12:**
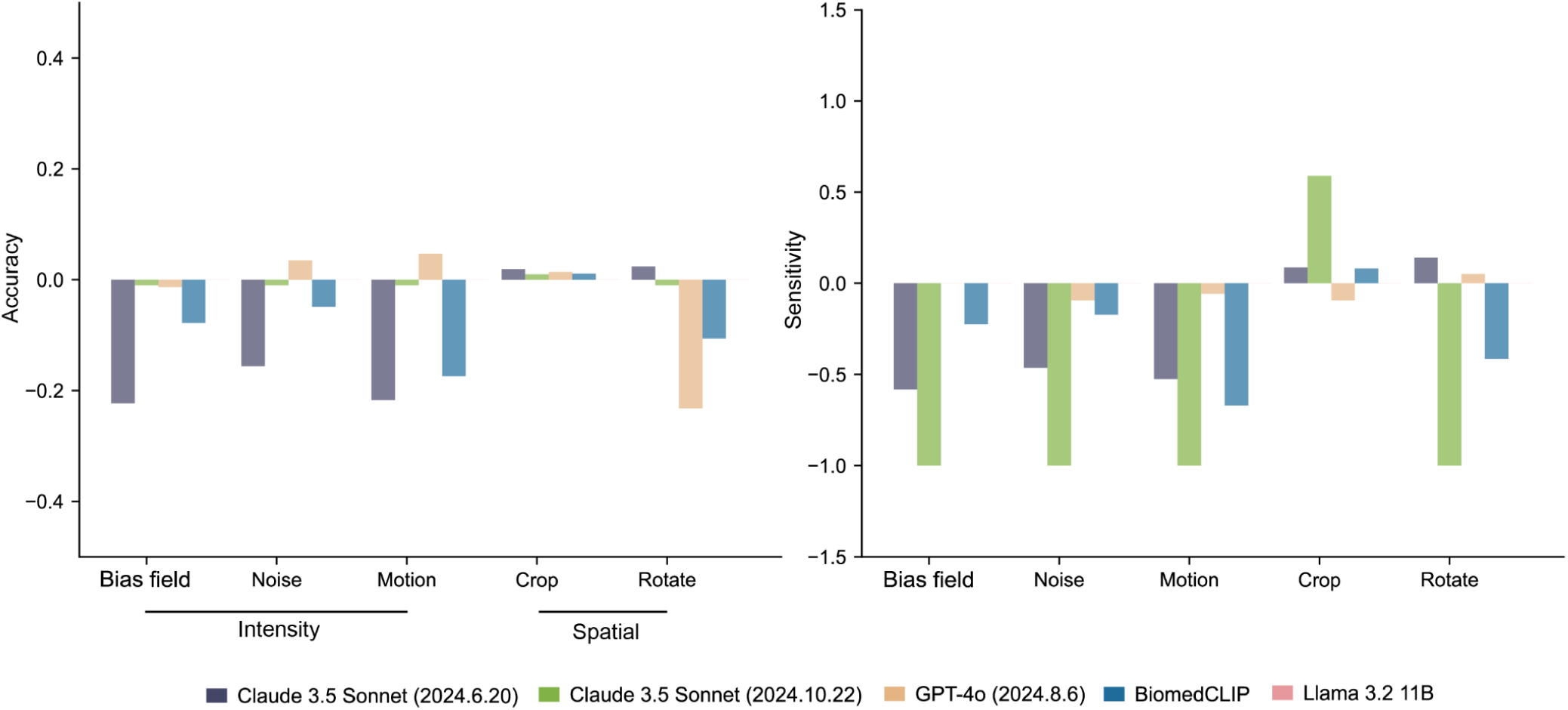
The performance percentage change of all Vision-Language models (VLMs) after adding weak artefacts to original OCT images using structured output prompts. Performance percentage change due to adding weak artefacts to original unaltered images are represented as y-axis. Complete quantitative data is available in Supplementary Table 8. For each task, we calculated performance percentage change through 1,000 iterations of stratified bootstrapping, with bar height representing mean value.

**Extended Data Fig. 13:**
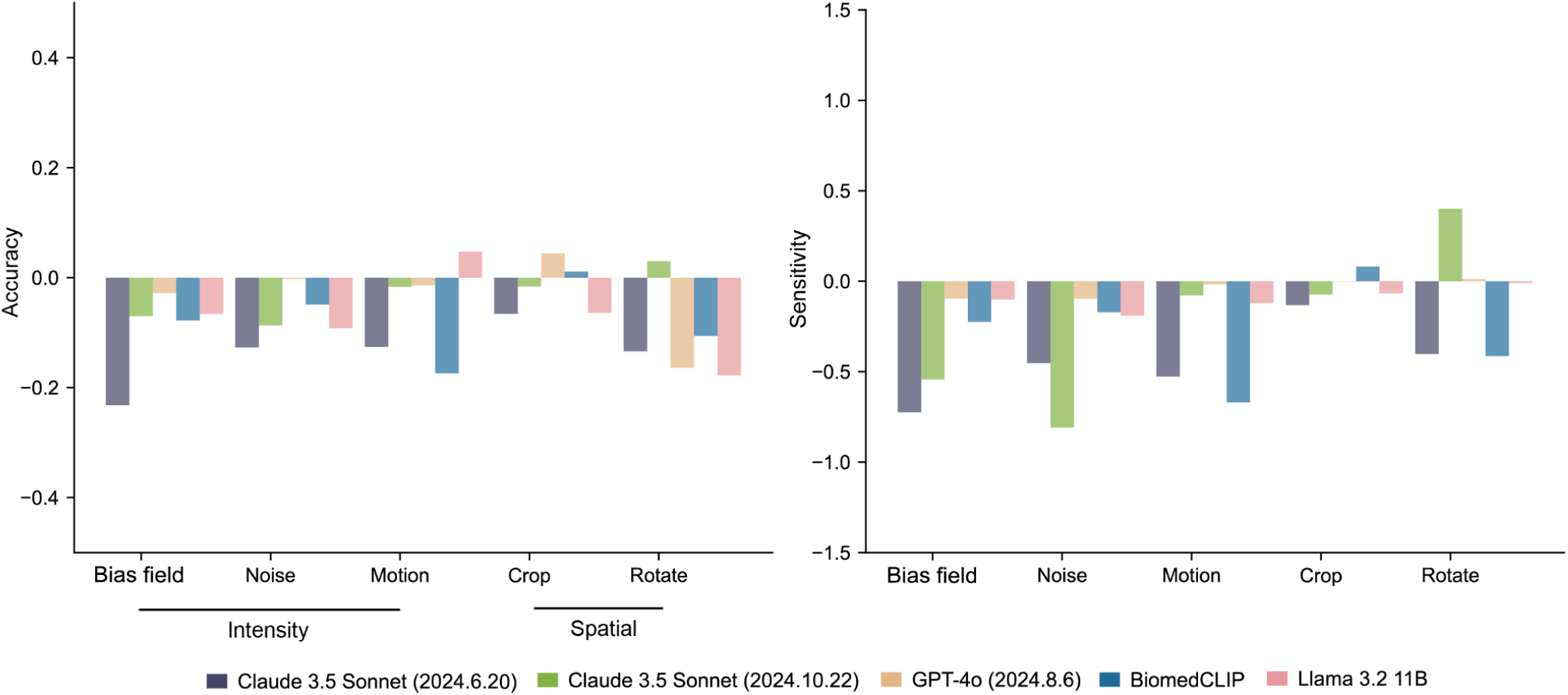
The performance percentage change of all Vision-Language models (VLMs) after adding weak artefacts to original OCT images using standard prompts. Performance percentage change due to adding weak artefacts to original unaltered images are represented as y-axis. Complete quantitative data is available in Supplementary Table 8. For each task, we calculated performance percentage change through 1,000 iterations of stratified bootstrapping, with bar height representing mean value.

**Extended Data Fig. 14:**
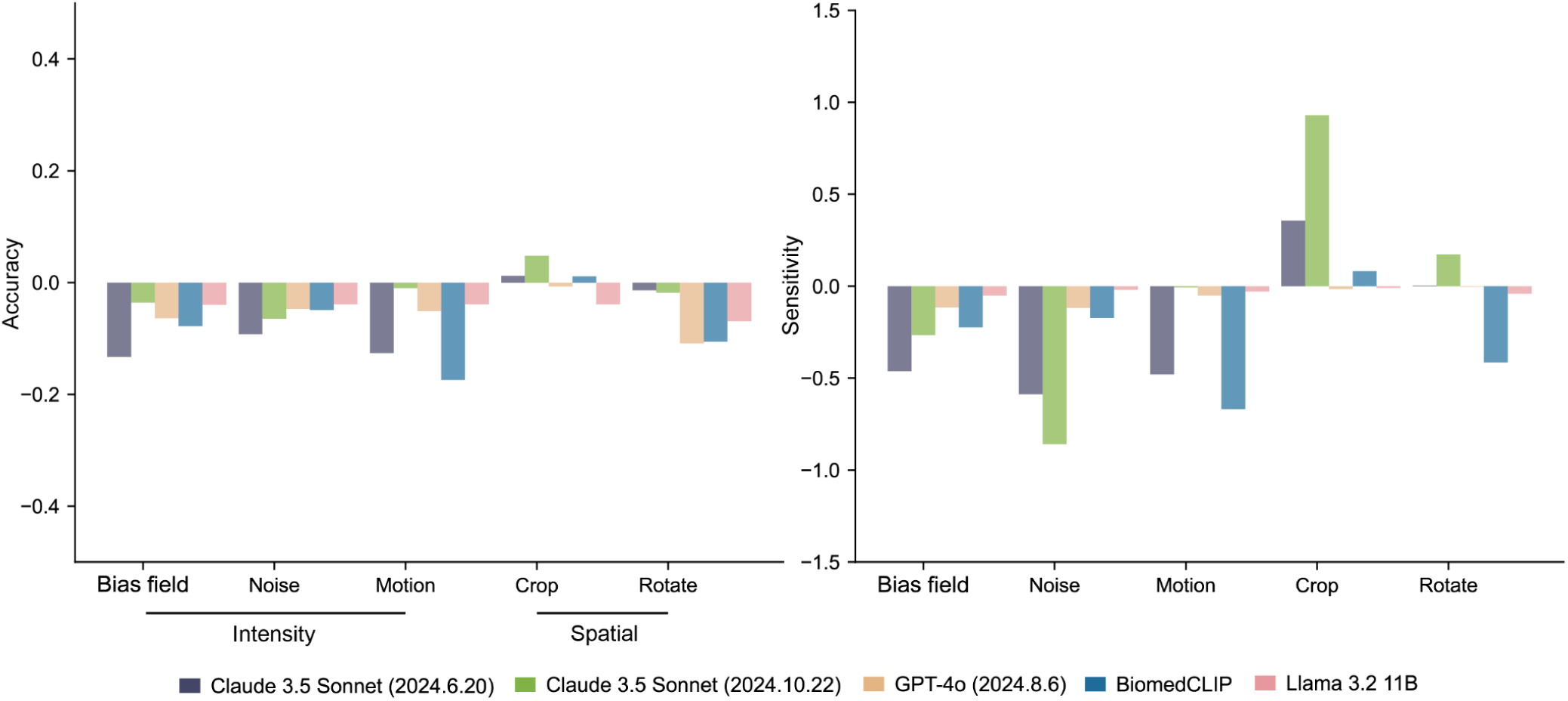
The performance percentage change of all Vision-Language models (VLMs) after adding weak artefacts to original OCT images using Chain of Thought prompts. Performance percentage change due to adding weak artefacts to original unaltered images are represented as y-axis. Complete quantitative data is available in Supplementary Table 8. For each task, we calculated performance percentage change through 1,000 iterations of stratified bootstrapping, with bar height representing mean value.

**Extended Data Fig. 15:**
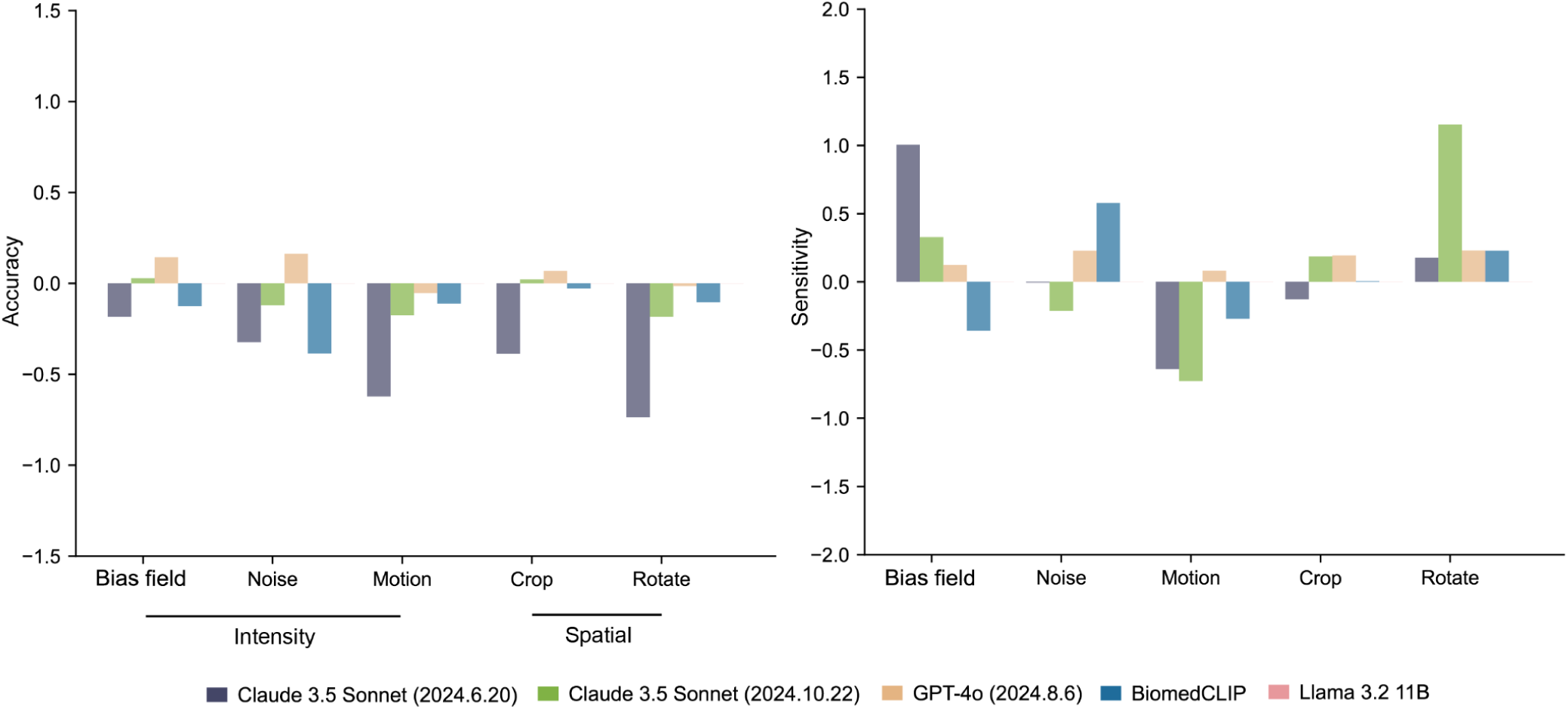
The performance percentage change of all Vision-Language models (VLMs) after adding weak artefacts to original X-ray images using structured output prompts. Performance percentage change due to adding weak artefacts to original unaltered images are represented as y-axis. Complete quantitative data is available in Supplementary Table 8. For each task, we calculated performance percentage change through 1,000 iterations of stratified bootstrapping, with bar height representing mean value.

**Extended Data Fig. 16:**
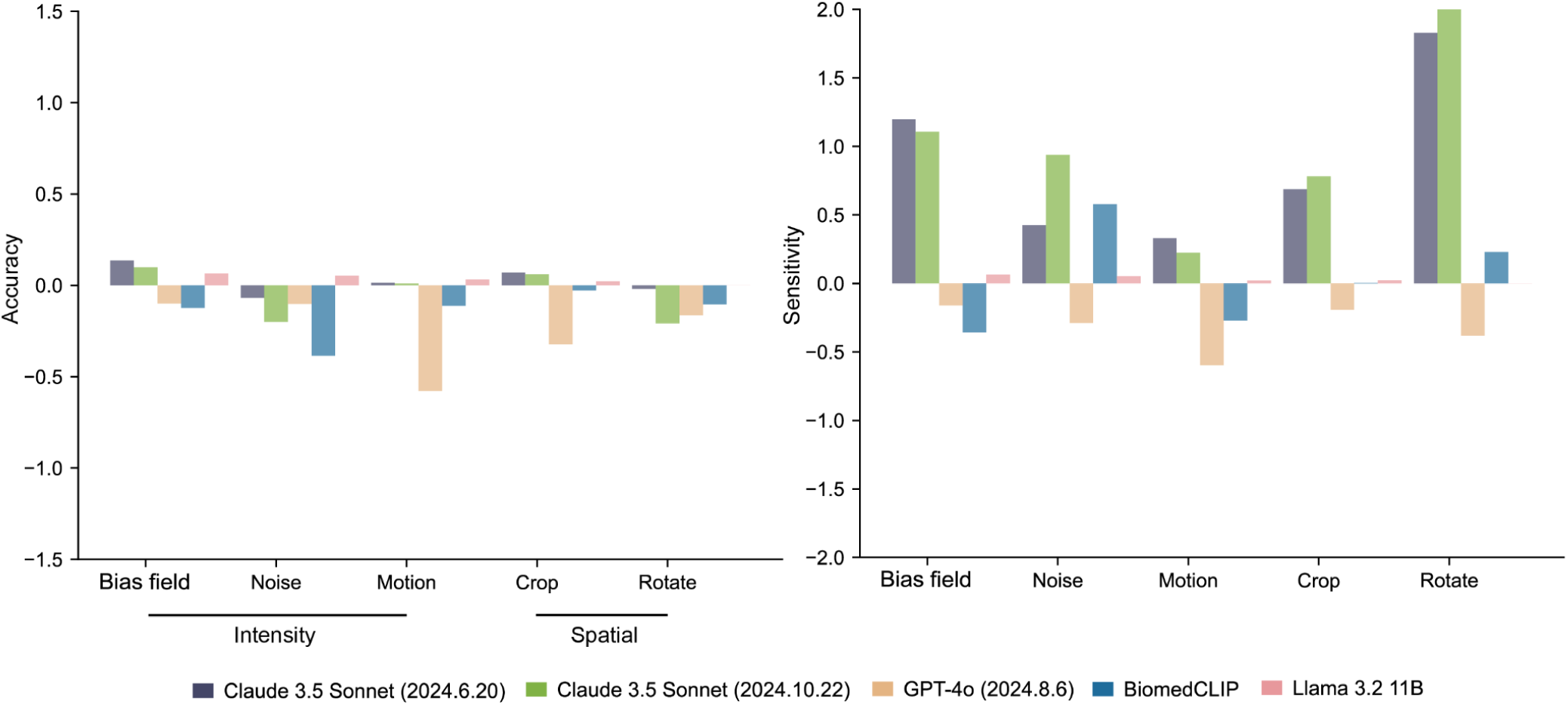
The performance percentage change of all Vision-Language models (VLMs) after adding weak artefacts to original X-ray images using standard prompts. Performance percentage change due to adding weak artefacts to original unaltered images are represented as y-axis. Complete quantitative data is available in Supplementary Table 8. For each task, we calculated performance percentage change through 1,000 iterations of stratified bootstrapping, with bar height representing mean value.

**Extended Data Fig. 17:**
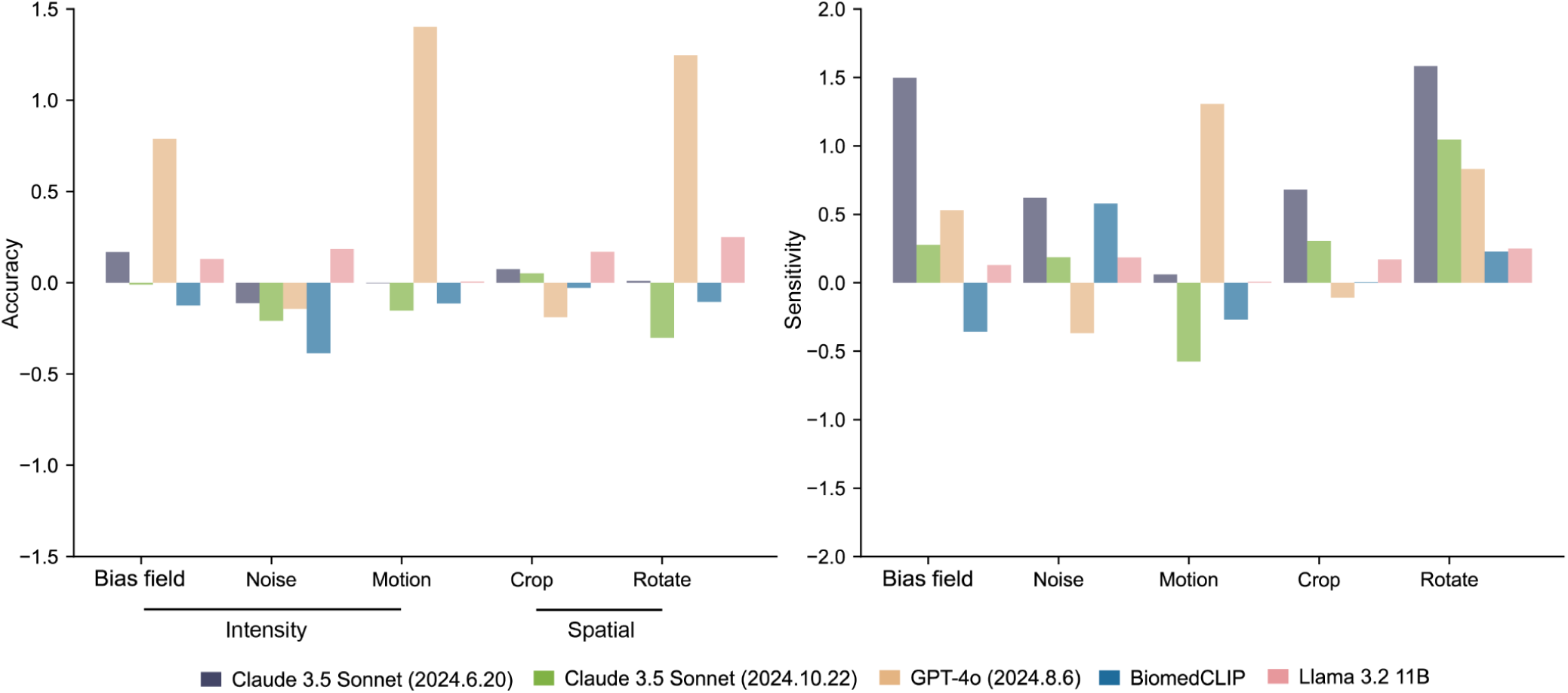
The performance percentage change of all Vision-Language models (VLMs) after adding weak artefacts to original X-ray images using Chain of Thought prompts. Performance percentage change due to adding weak artefacts to original unaltered images are represented as y-axis. Complete quantitative data is available in Supplementary Table 8. For each task, we calculated performance percentage change through 1,000 iterations of stratified bootstrapping, with bar height representing mean value.

**Extended Data Fig. 18:**
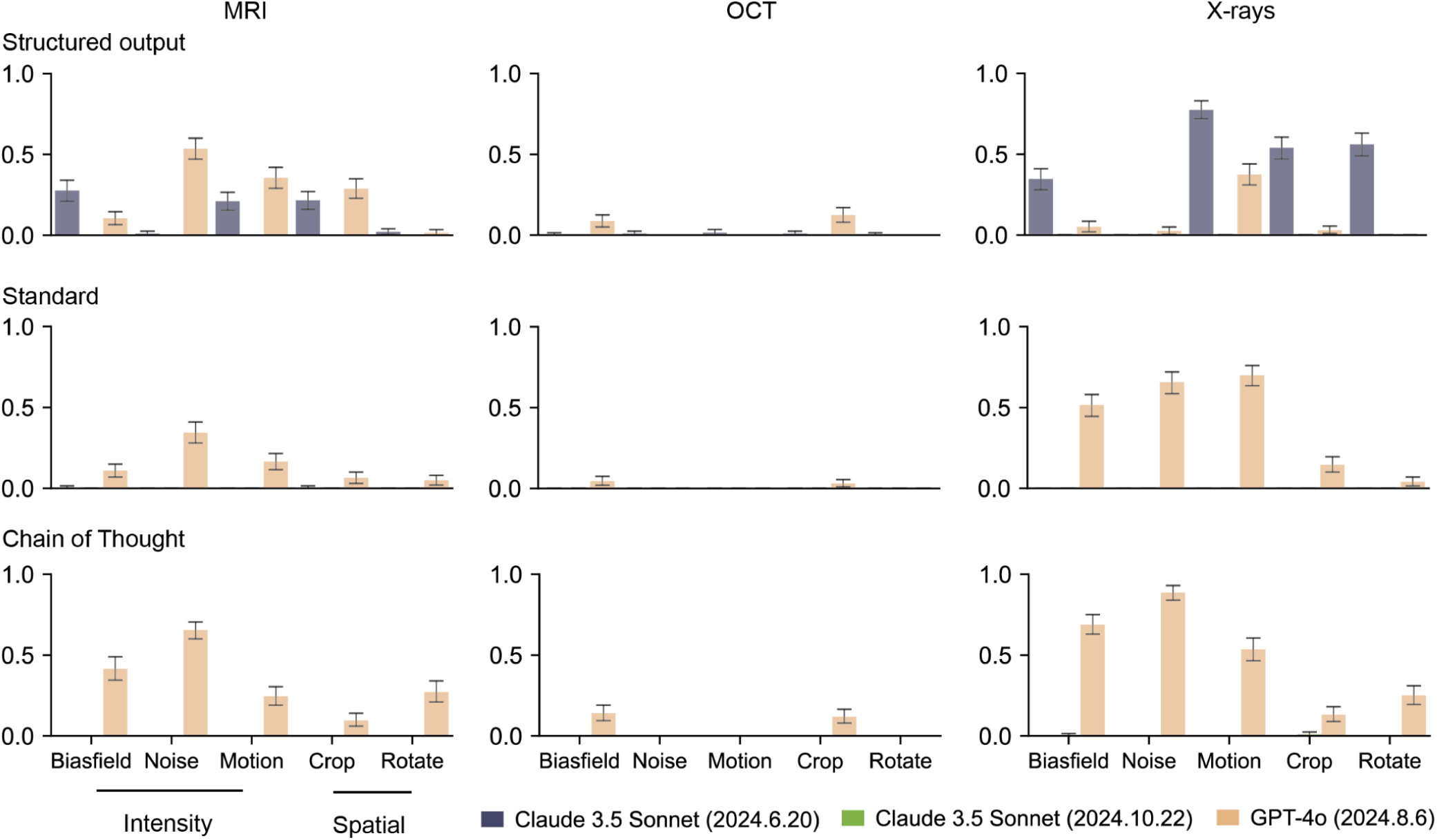
Vision-Language models (VLMs)’ refuse to answer rate after adding strong artefacts to the original unaltered images. The rate that VLMs refuse to answer due to ethical concerns or suggestions to consult specialists is shown in the y-axis. We excluded Llama due to poor accuracy with original unaltered images (Fig. 3 and Extended Data Fige. 4, 5) and BiomedCLIP because it only outputs probability. Quantitative results are presented in Supplementary Table 10. Results show mean detection rates with 95% confidence intervals (error bars).

**Extended Data Fig. 19:**
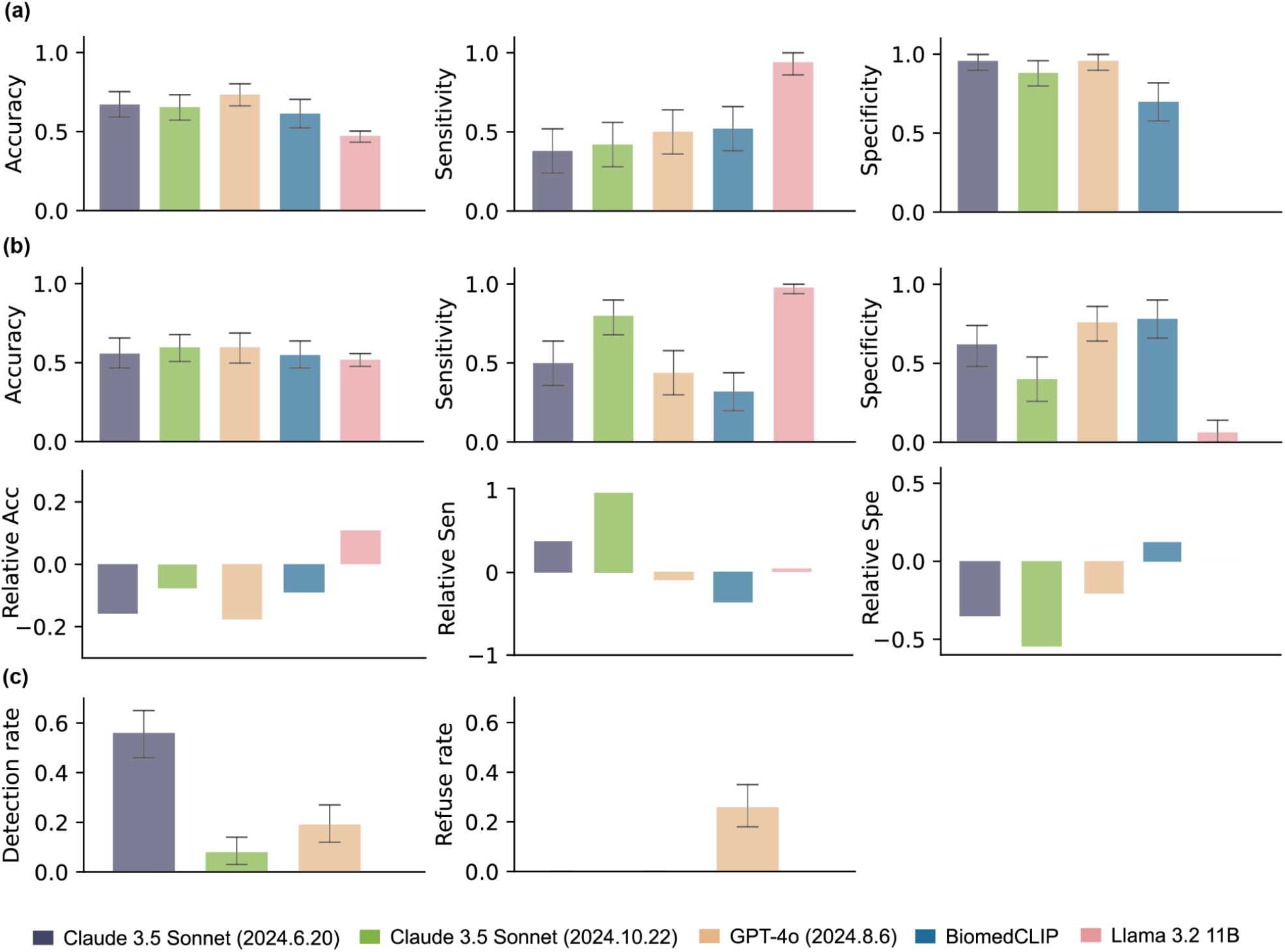
Robustness and strong artefact detection capability of Vision-Language Models (VLMs) in evaluating medical images with real-world artefacts. Figure (a) illustrates the models’ performance when evaluating high-quality medical images. Figure (b) presents the models’ performance on images with weak artefacts, along with their relative performance compared to high-quality images. Figure (c) displays the Vision-Language Models’ (VLMs) artefact detection rate and the refusal-to-answer rate when evaluating ungradable images. Llama is excluded due to its poor accuracy on high-quality images, and BiomedCLIP is excluded as it only outputs probabilities. Quantitative results are provided in Supplementary Table 11. The results show mean detection rates along with 95% confidence intervals (error bars).

